# Estimates and determinants of SARS-CoV-2 seroprevalence and infection fatality ratio using latent class analysis: the population-based Tirschenreuth study in the hardest-hit German county in spring 2020

**DOI:** 10.1101/2021.03.29.21254343

**Authors:** Ralf Wagner, David Peterhoff, Stephanie Beileke, Felix Guenther, Melanie Berr, Sebastian Einhauser, Anja Schütz, Hans Helmut Niller, Philipp Steininger, Antje Knöll, Matthias Tenbusch, Clara Maier, Klaus Korn, Klaus J. Stark, Andre Gessner, Ralph Burkhardt, Michael Kabesch, Holger Schedl, Helmut Küchenhoff, Annette B. Pfahlberg, Iris M. Heid, Olaf Gefeller, Klaus Überla

## Abstract

SARS-CoV-2 infection fatality ratios (IFR) remain controversially discussed with implications for political measures, but the number of registered infections depends on testing strategies and deduced case fatality ratios (CFR) are poor proxies for IFR. The German county of Tirschenreuth suffered a severe SARS-CoV-2 outbreak in spring 2020 with particularly high CFR.

To estimate seroprevalence, dark figure, and IFR for the Tirschenreuth population aged ≥14 years in June/July 2020 with misclassification error control, we conducted a population-based study, including home visits for elderly, and analyzed 4203 participants for SARS-CoV-2 antibodies via three antibody tests (64% of our random sample). Latent class analysis yielded 8.6% standardized county-wide seroprevalence, dark figure factor 5.0, and 2.5% overall IFR. Seroprevalence was two-fold higher among medical workers and one third among current smokers with similar proportions of registered infections. While seroprevalence did not show an age-trend, the dark figure was 12.2 in the young versus 1.7 for ≥85-year-old. Age-specific IFRs were <0.5% below 60 years of age, 1.0% for age 60-69, 13.2% for age 70+, confirming a previously reported age-model for IFR. Senior care homes accounted for 45% of COVID-19-related deaths, reflected by an IFR of 7.5% among individuals aged 70+ and an overall IFR of 1.4% when excluding senior care home residents from our computation.

Our data underscore senior care home infections as key determinant of IFR additionally to age, insufficient targeted testing in the young, and the need for further investigations on behavioral or molecular causes of the fewer infections among current smokers.

## Introduction

COVID-19 case numbers continue to rise world-wide, but the precise cumulative number of cases remains unknown. Testing frequencies and strategies vary largely between countries and over time limiting the strength of the conclusions that can be drawn. The large percentage of asymptomatic infections further reduces the percentage of SARS-CoV-2 infections registered [1]. Not knowing the total number of SARS-CoV- 2 infections also impairs the calculation of infection fatality ratios (IFRs) and the level of herd immunity. Determining the ratio of non-registered to registered SARS-CoV-2 infections, the dark figure, helps understand the extent of undetected infections. The gold standard for assessing total cumulative case numbers for viral infections are population-based seroprevalence studies within an appropriate time-period after outbreak based on random sampling from public registries. At low seroprevalence, even a small deviation from 100% specificity of the tests used for determining antibody responses to SARS-CoV-2 can lead to bias due to a low positive predictive value. Additional adjustments considering the decay of antibody levels after infection may also be necessary, although recent results indicate stability of IgG levels against the SARS-CoV-2 spike protein for more than six months [2].

In Europe, Italy was the first country to be hit hard with more than 1 confirmed COVID-19 case / 100 000 inhabitants / 14 days nationwide on 28^th^ of February, 2020. With a delay of one to two weeks, case counts in other European countries, such as Spain, France, Germany, UK and Portugal passed this level[3,4]. In Germany, a first cluster of COVID-19 cases occurred between January 27^th^ to February 19th, 2020, but was contained by contact tracing[5,6]. Most likely, ski-travelers returning from Austria and Italy and the carnival festivities were important determinants of the subsequent initial spread of SARS-CoV-2 in Germany and the number of confirmed cases exceeded 1 000 by March 10^th^. The consequence of a superspreading event during carnival on February 15^th^ in Gangelt, a municipality in the county of Heinsberg in North Rhine Westphalia, was analyzed by a household-based, seroprevalence study. While 3.1% of the population had been reported SARS-CoV-2 positive by PCR at the time of the study, the seroprevalence at this time in Gangelt was 14.11%. Based on only 7 early deaths of COVID-19 cases reported until April 6^th^ in Gangelt, this resulted in an inferred case-fatality ratio of 1.8% and an inferred infection fatality ratio of 0.36% [7].

A second hotspot of COVID-19 cases in Germany occurred in the county of Tirschenreuth located in the North- East of Bavaria with the first recognized COVID-19 case on February 17^th^, 2020. Daily cases counts peaked at 55 on March 16^th^ and a stay-at-home order was issued for the hardest hit municipality in that county on March 18^th^. Until May, 11^th^ the incidence of new COVID-19 cases declined to sporadic cases. By then, the number of total cases had summed up to 1122 corresponding to 1548 cases/100,000 inhabitants[8,9], the highest incidence observed in Germany for any county until November 2020. In addition, an extraordinary high case-fatality ratio (CFR, 11.5%) prompted an epidemiological investigation by the Robert Koch Institute, Germanýs federal center for infectious disease control. This investigation concluded that travelers returning from skiing vacations in Italy or Austria prior to detection of the first case in Tirschenreuth, early undetected community transmissions, and ultimately a beer festival contributed to the steep increase in COVID-19 cases starting March 10^th^, but were not sufficient to explain it entirely[8,9].

One explanation for the high CFR could be an increased infection occurrence in the most vulnerable age groups: median age of COVID-19 cases in the county of Tirschenreuth was 56 years compared to Germany and Bavaria (50 years each) or Gangelt (52 years) and the percentage of COVID-19 cases aged 70 years or older among all COVID cases was higher in Tirschenreuth than in the rest of Germany [9]. In addition, 56 of the 129 deaths observed in Tirschenreuth until May 11^th^ occurred in senior care homes resulting in CFRs of 36% for residents of senior care homes and 7.5% for inhabitants of Tirschenreuth not living in care homes [9]. However, both CFRs are higher than the ones observed for residents of care homes for the elderly (20%) or the general population (4.4%) in Germany during the same time period. In addition, the CFR in the general population in the county of Tirschenreuth is more than 4-fold higher than the CFR initially reported for the municipality of Gangelt [7].

The small percentage of asymptomatic cases among the registered cases in Tirschenreuth [9] further suggests that a larger proportion of asymptomatic SARS-CoV-2 infections remained undetected, particularly at this very early phase of the pandemic in Europe. Since difference in testing frequencies and strategies may affect the percentage of reported SARS-CoV-2 infections and thus the CFR, the determination of the seroprevalence rate in the Tirschenreuth population will allow to calculate the IFR and help to better understand the discrepant observations regarding the CFR.

The IFR is an important parameter to predict the risk that health care systems are overwhelmed by the COVID-19 pandemic. Many political decisions on contact reduction measures are still made without a good knowledge on the precise IFR for the targeted population. A recent meta-analysis also revealed that the IFR can vary substantially across different locations [10] possibly reflecting differences in age and risk structure among others.

To determine the seroprevalence, the dark figure and the IFR in the county of Tirschenreuth, we determined the prevalence of antibodies against SARS-CoV-2 in a population-based study based on a random sampling approach and three independent SARS-CoV-2 antibody tests. Demographic and lifestyle factors potentially associated with seroprevalence were also assessed by a questionnaire.

## Subjects and Methods

### Study design

The county Tirschenreuth located in the north-eastern part of Bavaria immediately adjacent to the Czech Republic is known as one of the hot spots of SARS-CoV2 spread in Germany during spring and early summer 2020 with - until November 2020 - the highest percentage in PCR diagnosed infections / 100 000 inhabitants all over Germany until November 2020.

TiKoCo was designed as prospective cohort study with a baseline survey to determine the SARS-CoV- 2 seroprevalence in the county of Tirschenreuth and two follow-up investigations 4 and 9 months after baseline to monitor its temporal development focusing on the durability of antibody responses, incidence of new infections and potential secondary infections. Further primary goals of this baseline survey are to quantify the percentage of undetected infections (“dark figure”) and to determine the infection fatality ratio (IFR) in the overall population and depending on age, sex and residency in the various municipalities of the county Tirschenreuth. We focused to yield robust data on seroprevalence by determining serum antibodies directed against two different viral antigens, the SARS-CoV-2 nucleoprotein as well as the viral spike protein (i) (S) and the S protein receptor-binding domain (RBD) by 3 independent assays. Further aspects of interest were (i) the association of the individuals’ report of COVID-19 related symptoms and antibody status, particularly among these who were not aware of their infection (i.e. excluding individuals with a report of positive PCR-test) and (ii) the association of education, individual household situation and lifestyle factors with antibody status.

The TiKoCo study was approved by the Ethics Committee of the University of Regensburg, Germany (vote 12-101-0258) and adopted by the Ethics Committee of the University of Erlangen (vote 248_20 Bc). The study complies with the 1964 Helsinki declaration and its later amendments. All participants provided written informed consent.

### Study capture area and eligible individuals

The county Tirschenreuth captures 64 643 aged 14 years or older inhabitants of mostly Caucasian ethnicity including 12 066 aged 70+. The county comprises 26 municipalities with populations varying from 822 to 7 807, respectively. Eligibility criteria for participation was the willingness to come to the indicated study centre and to provide 5.7 ml blood. Inclusion criteria were legal age and univalent consent of custodial parents for those aged < 18 years, German language skills sufficiently good to understand the participant information and to respond to the questionnaire. Children below 14 years and individuals with legal guardian were excluded from participation.

### Sample size

Assuming a SARS-CoV-2 seroprevalence of 10% in the population of the county Tirschenreuth, the participation of 3 600 individuals would result in 95% confidence intervals with a width of +/- 1%, which was considered satisfactory. With regard to a future longitudinal survey, also changes in seroprevalence between two sampling time points of 0.1% could be proven statistically significant at a predefined statistical power of 80%. We were conservatively assuming a response rate of 50% or above for our study. This suggested that up to 7 200 randomly sampled individuals were to be contacted to successfully recruit the calculated 3 600 baseline study participants.

### Study participant recruitment

For the baseline survey, we selected a representative sample of 7 200 residents of the Tirschenreuth county aged 14 years and older by means of a sex- and municipality-stratified random sample. Based on data on county residents regarding sex and age, which were available for each municipality (Bayerisches Landesamt für Statistik 2019), we determined, for each of the 26 municipalities, the number of men and women, which corresponded to the share of the members of the municipality within the total population of the county Tirschenreuth, respectively.

The various municipalities in the county Tirschenreuth were approached with specific target figures and asked to randomly sample men and women aged 14 years or older (as per June 2^nd^), collectively adding up to a total sample of 7 200 potential study participants (**Figure 1**). Between June 17^th^ and June 26^th^, we mailed invitation letters to two thirds of these pre-selected individuals with the request to participate in a prospective COVID-19 antibody study. Invitations included written information regarding the study plan and goals, participant information, and a questionnaire (see below). Depending on their home address, invited individuals were asked to visit the nearest of the three study centers for a given date between June 28^th^ and July 10^th^ to donate 5.7 ml blood, to deliver the filled-in questionnaire, receive the data protection declaration, obtain personal advice from study staff if requested and sign the declaration of consent. 104 handicapped or otherwise immobile individuals were offered a visit at home for the blood draw. Invited individuals with flu-like symptoms were asked to stay at home and use the installed hotline to agree on a date for a visit at home for blood draw, additionally offering a nasal throat swab. Changing the date of and location for the blood sampling via the hotline was optional. After the 2^nd^ day of blood sampling (June 29^th^), the participation ratio based on the 4801 initially contacted persons was calculated to decide on the number of individuals for the second batch of invitations. This resulted in another 1 807 individuals who were contacted via mail and asked to visit one of the test centers in the week of July 13^th^. Altogether 6540 individuals were contactable.

**Figure 1.**
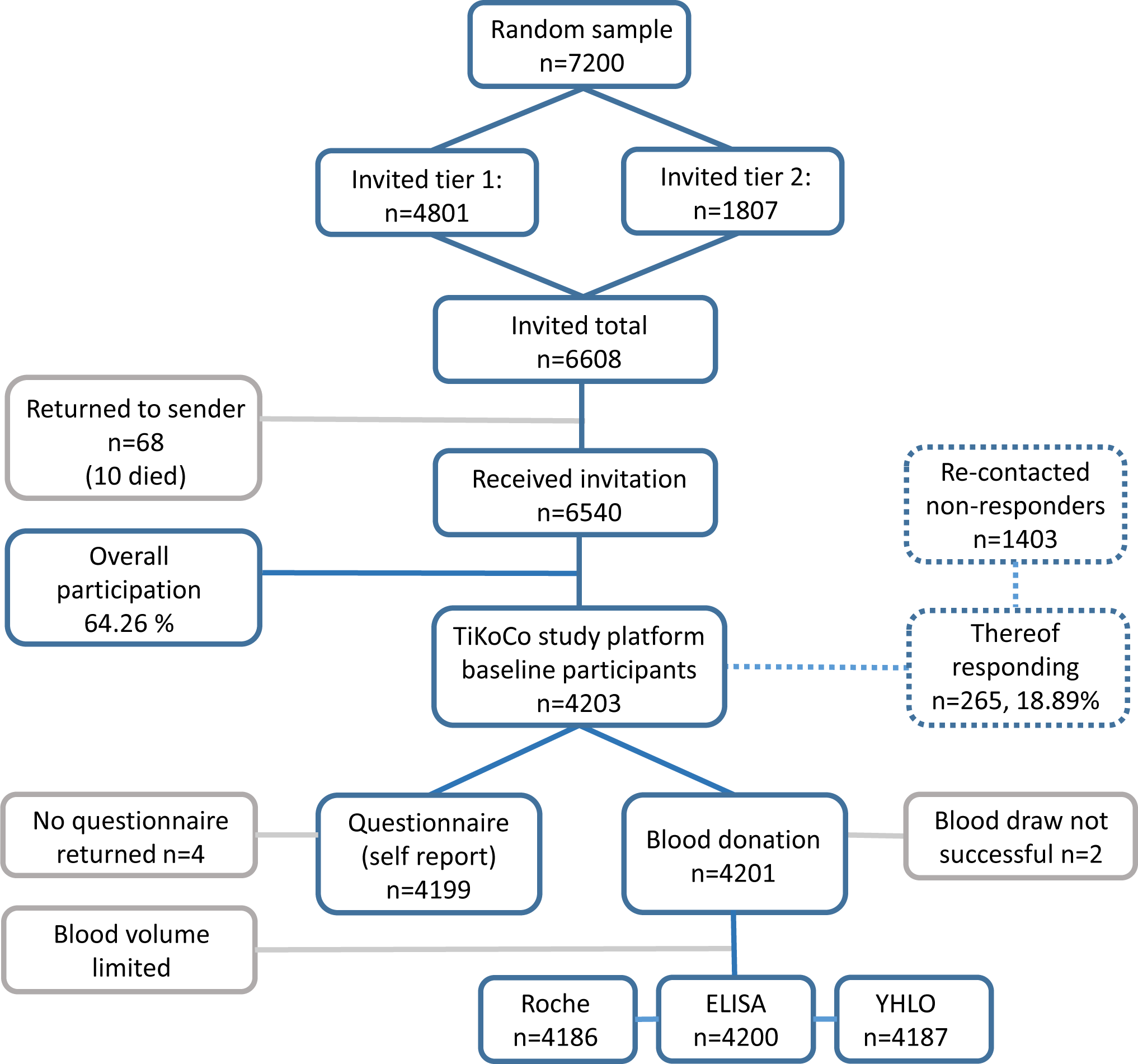
Summary of the TiCoKo study design. The strategy and numbers (n) underlying the random sampling and recruitment of study participants, the collection of information via a self-reporting questionnaire as well as the testing strategy to determine the true serostatus of the participants via three independent test formats (Roche-Cobas, *in-house* ELISA and YHLO) is shown. Measures to minimize and understand non-response are indicated (dashed line), reasons for drop-outs of participants, numbers of returned questionnaires and successful blood samples are given (grey line). *Figure was designed using PowerPoint 365 for Windows, Microsoft, Redmond Washington USA,* www.microsoft.com

### Measures to minimize and understand non-response

Individuals who showed up at the study center at the dates as communicated in the invitation letter or as otherwise agreed via the hotline were considered immediate responders. A reminder invitation letter was sent out to the 1403 initial non-responders who were re-invited for July 13^th^ to July 17^th^. Those who responded following this reminder were considered late responders (n=265). Of the 6 608 sent out invitation letters, 68 were returned (not contactable individuals). It turned out that the residents at 10 of these addressees had died (reason unknown) and one was hospitalized due to COVID-19. Individuals who received the invitation and did not visit the study center or asked for an alternative appointment, not even after the invitation reminder, were considered as non-responder.

### Questionnaire

Together with the invitation mailed to individuals eligible for our study, we sent out a self-administered questionnaire and asked individuals to bring the filled-out questionnaire to the study center offering personal counseling by trained staff members in case of questions. The questionnaire was, in part, based on the questionnaire developed within the spring 2020 survey in the German National Cohort (kindly provided by the GNC) [11] and also utilized in the AugUR study [12]. This enables comparability of TiKoCo questionnaire results with GNC results for those aged 20-69 years and with AugUR for those aged 79 years and older.

In the questionnaire, we asked for (i) COVID-19 related symptoms (cough, shortness of breath, respiratory problems, fever, chills, loss of smell/taste, bronchitis/pneumonia), other symptoms related to general infections (pain in extremities, diarrhea, nausea, red eye/eye infection, headache, fatigue, rhinitis), (ii) PCR-based SARS-CoV-2 testing (results, date, reason, symptoms at time of testing), hospitalization related to COVID-19 or generally to bronchitis/pneumonia since the start of the pandemic (as per Feb 1st, 2020); (iii) household (living alone, ≥ 1 other person, nursing home); (iii) previous illnesses by self-report (heart disease, lung disease including asthma and chronic bronchitis, kidney disease, diabetes, hypertension, cancer, auto- immune disease, blood coagulation disorder); (iv) education (highest level of school, university and/or vocational training) and employment (status, type of occupation); (v) lifestyle factors: smoking (status as current, former, never; number of cigarettes smoked per day); alcohol consumption (frequency of drinking, number of drinks typically consumed); TV consumption (days per week with TV consumption for >2 hours); physical activity as categories of weekly hours of light activity (0, 0-2 hours, 2-4 hours, > 4 hours; including bicycling, walking).

### Blood draw, transport and antibody measurements

Blood was drawn in sitting position by qualified study personnel into a barcoded serum monovette (Sarstedt AG Co.KG, Nümbrecht, Germany). All tubes sampled per day were processed immediately. One aliquot was sent to University Hospital Erlangen for assessment in the YHLO SARS CoV-2 test (chemoluminescent immune assay, CLIA). Two tests, an in-house ELISA and Elecsys, Roche Diagnostics (Roche-Cobas, CLIA) were performed at the University Hospital Regensburg.

The Elecsys Anti-SARS-CoV-2 test (Roche Diagnostics GmbH, Penzberg, Germany) detecting nucleoprotein- (N)-directed complete Ig was operated on the Cobas pro e 801 module and the YHLO SARS CoV-2 test (Shenzhen Yhlo Biotech Co., Ltd., Shenzen, China) detects IgG antibodies to the N- and S-protein on the iFlash 1800 according to the manufacturers recommendations, respectively. A validated in-house ELISA detecting IgG antibody responses to the receptor binding domain (RBD) of the SARS-CoV-2 spike protein was performed essentially as described earlier [13].

### Latent class analysis to derive the true seropositivity status and seroprevalence in the study population

We used three different antibody tests. As there is no gold standard for defining the true status, we used a latent class analysis (LCA) to define the true seropositivity status based on the pattern of results from all three antibody tests. LCA is a classical statistical methodology to identify a set of discrete, mutually exclusive latent (i.e. unobserved) classes based on the observed pattern of a set of categorical variables. LCA has been introduced by Lazarsfeld [14] and has been repeatedly used in the context of defining the true status of a disease when a gold standard procedure for its detection is missing [15]. The basic idea of LCA in our setting is to treat the unobservable true seropositivity status as being equivalent to two latent classes and to relate the observed antibody test results from the three tests to it via a statistical model. The critical model assumption is that the three antibody tests are independent conditional on the true seropositivity status. The validity of this assumption can be tested with different methods, we used the log-odds ratio check proposed by Garrett and Zeger [16].

Application of LCA to our data yields a statistical prediction of the true seropositivity status for each study subject. This allows, in a subsequent step, the estimation of seroprevalence in the study population and its subgroups defined by gender, age and residence in local municipalities of the county of Tirschenreuth. Point estimates of seroprevalence are always accompanied by asymptotic 95%-confidence intervals (CI), computed by Wilson’s method, to provide the information of their precision. The statistical software SAS version 9.4 (SAS Institute Inc., Cary, North Carolina, USA) has been used for these computations, LCA modeling has been performed employing a SAS extension [17].

### Standardization to derive seroprevalence in study capture area

The statistical analysis of seropositivity data in our study sample yields crude results on seroprevalence and associated measure like dark figure factors and infection fatality rates applicable to the study cohort only. Since our study sample represents a random sample of residents of the Tirschenreuth county aged 14 years and older, we were able to compute standardized figures of these quantities that allow a population-oriented interpretation for the Tirschenreuth county. We used official information from the administrative county office of Tirschenreuth and the 26 local residents’ registration offices in the county of Tirschenreuth to derive its age and sex distribution as well as the distribution of the population over the municipalities of the county. By weighting and combining the age-, sex- and the municipality-specific figures estimated from the study sample with their corresponding weights in the Tirschenreuth population we were able to derive seroprevalences and associated measures standardized for all three factors (in situations when we report subgroup-specific results, we standardized for the two factors not defining the subgroup).

When addressing the effect of residence in senior care homes, we divided the municipalities of the county into two subgroups: one group of municipalities with at least one senior care home and the remaining group of municipalities without any senior care home. In a further step, we also calculated standardized seroprevalence, dark figure factors and IFR for the individuals of all those above 70 years of age not residing in a senior care home and for individuals across all age groups not residing in a senior care home (in this additional analysis, the 13 study participants residing in a senior care home were excluded when computing crude seroprevalences and the 920 inhabitants residing in senior care home in the county of Tirschenreuth were excluded when computing the weights for the standardization under the assumption that their age distribution resembles the age distribution of senior care inhabitants in Germany [18].

All 95%-CIs for standardized seroprevalences were computed using Wilson’s method for binomial proportions assuming the weights as fixed constants. For dark figure factors and infection fatality ratios, simple CIs for binomial proportions ignore the uncertainty in officially reported infection and death counts. Therefore, we do not report such intervals for these figures; instead, we computed Bayesian credibility intervals accounting for these uncertainties in the following manner: for the upper and lower bound of interval, we used the empirical 2.5% and 97.5% quantiles of 100 000 samples drawn from a beta distribution with parameters dependent on the observed counts of infections and deaths, respectively, where in each of the 100 000 samples the seroprevalence incorporated in the calculation was sampled from a normal distribution with mean and standard deviation given by the standardized estimates in the corresponding study (sub-)group. This approach follows the method applied by Streeck et al. [7].

### Statistical models to investigate the association of seropositivity with potential risk factors

We used logistic regression to investigate the association of seropositivity (binary, LCA-derived) with potential risk factors. We considered the following person-specific covariates: 1) person characteristics age group (14-19, 20-69, 70+) and sex; 2) personal background as education in years (6-10, 11-15, 16-23), household size (1, 2, 3-5, 6+) and occupation during spring 2020 (grocery store, medicine, other), and 3) lifestyle factors as physical activity (high, low), alcohol consumption (drinks per day: 0, 0-0.25, 0.25-1, >1), body-mass-index (<18.5, 18.5-25, 25-30, >30), and smoking (current, ex, never). To further investigate the association of smoking with seropositivity, we performed additional analyses on the dose-response association of seropositivity with the number of smoked cigarettes (linear or non-linear, in the overall sample and current smokers, adjusted for age and sex). The associations were presented as odds ratios with 95%- profile likelihood confidence intervals. Associations with a p-value <0.05 were considered as significant

### Comparison of reported symptoms between persons with and without SARS-CoV-2 infection

We analyzed the self-reported information on occurrence of COVID-19 related symptoms since beginning of the pandemic by comparing the fraction of individuals reporting the specific symptoms between three groups: seronegatives, seropositives with self-reported positive PCR test (i.e., individuals aware of their infection), and seropositives without positive PCR test (individuals unaware of infection). Furthermore, we quantified the association of symptoms with seropositivity based on odds ratios of symptom occurrence and seropositivity combining individuals with and without positive PCR test result in the sero-positive group.

## Results

### Participant characteristics and response

Of the Tirschenreuth inhabitants with ≥ 14 years of age, 6 608 individuals were randomly selected in a sex-, and municipality-stratified fashion and invited to participate in the TiKoCo study in June 2020 and 6 540 individuals obtained the invitation letter. Of the contactable individuals, 4203 individuals participated yielding an overall net response of 64.26% (**Figure 1**). The response was higher among the age group 20-69 compared to those 14-19 years or 70+ years of age (**Supplemental Figure 1**) and also differed amongst the 26 local municipalities (**Supplemental Table 1**). The 4 203 participants included 48.3% men, age ranged from 14 to 102 years (**Table 1**). 20.4% reported to be current smokers and median body-mass-index was 27.1 kg/m² and 26.0 kg/m² among male and female participants, respectively. When focusing on the 633 participants aged 70+, which is the age group most at risk for severe COVID-19 and COVID-19 related death, 7.2% were current smokers smoking on average 11 cigarettes per day and median BMI was 27.5 kg/m², 1.9% (n=15) lived in a nursing home. When comparing our participants aged 70+ to the population-based AugUR study, which focuses per design on elderly aged 70+, we found no major differences in demographic or lifestyle factors, except for a higher proportion of current smokers and a higher number of daily smoked cigarettes (AugUR 4.8% current smokers, average of 6 cigarettes per day).

**Table 1.**
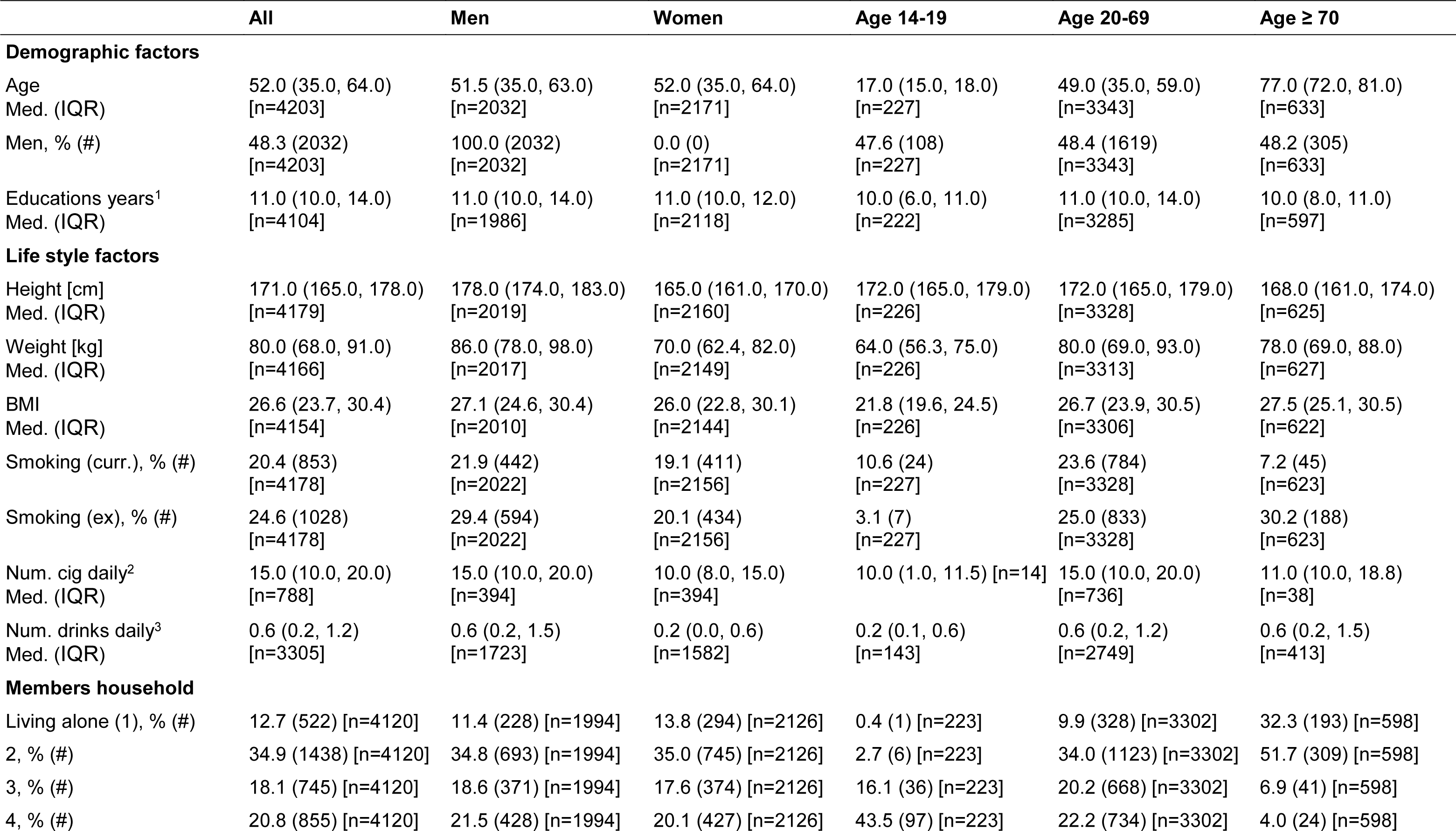
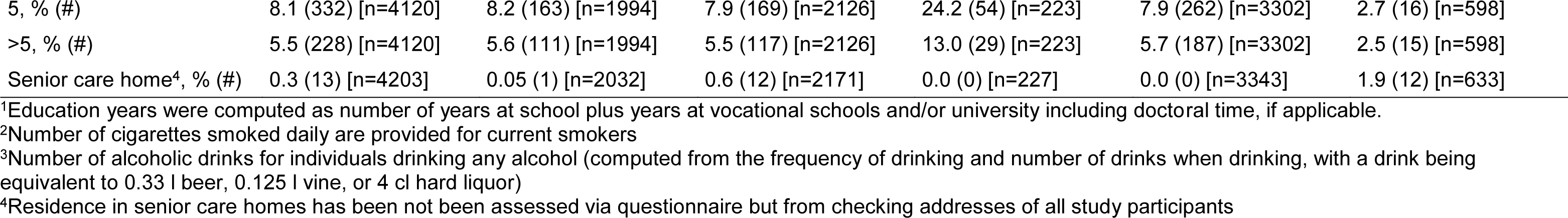
Participant characteristics. Shown are characteristics of the 4 203 study participants (age, sex) and other characteristics for the 4201 study participants who filled out the questionnaire. Shown are median and (interquartile-range, IQR) or relative (%) and absolute (#) frequencies compared to the non-missing [number of individuals the category].

|  | All | Men | Women | Age 14-19 | Age 20-69 | Age ≥ 70 |
| --- | --- | --- | --- | --- | --- | --- |
| **Demographic factors** |  |  |  |  |  |  |
| Age | 52.0 (35.0, 64.0) | 51.5 (35.0, 63.0) | 52.0 (35.0, 64.0) | 17.0 (15.0, 18.0) | 49.0 (35.0, 59.0) | 77.0 (72.0, 81.0) |
| Med. (IQR) | [n=4203] | [n=2032] | [n=2171] | [n=227] | [n=3343] | [n=633] |
| Men, % (#) | 48.3 (2032) | 100.0 (2032) | 0.0 (0) | 47.6 (108) | 48.4 (1619) | 48.2 (305) |
|  | [n=4203] | [n=2032] | [n=2171] | [n=227] | [n=3343] | [n=633] |
| Educations years<sup>1</sup> | 11.0 (10.0, 14.0) | 11.0 (10.0, 14.0) | 11.0 (10.0, 12.0) | 10.0 (6.0, 11.0) | 11.0 (10.0, 14.0) | 10.0 (8.0, 11.0) |
| Med. (IQR) | [n=4104] | [n=1986] | [n=2118] | [n=222] | [n=3285] | [n=597] |
| **Life style factors** |  |  |  |  |  |  |
| Height [cm] | 171.0 (165.0, 178.0) | 178.0 (174.0, 183.0) | 165.0 (161.0, 170.0) | 172.0 (165.0, 179.0) | 172.0 (165.0, 179.0) | 168.0 (161.0, 174.0) |
| Med. (IQR) | [n=4179] | [n=2019] | [n=2160] | [n=226] | [n=3328] | [n=625] |
| Weight [kg] | 80.0 (68.0, 91.0) | 86.0 (78.0, 98.0) | 70.0 (62.4, 82.0) | 64.0 (56.3, 75.0) | 80.0 (69.0, 93.0) | 78.0 (69.0, 88.0) |
| Med. (IQR) | [n=4166] | [n=2017] | [n=2149] | [n=226] | [n=3313] | [n=627] |
| BMI | 26.6 (23.7, 30.4) | 27.1 (24.6, 30.4) | 26.0 (22.8, 30.1) | 21.8 (19.6, 24.5) | 26.7 (23.9, 30.5) | 27.5 (25.1, 30.5) |
| Med. (IQR) | [n=4154] | [n=2010] | [n=2144] | [n=226] | [n=3306] | [n=622] |
| Smoking (curr.), % (#) | 20.4 (853) | 21.9 (442) | 19.1 (411) | 10.6 (24) | 23.6 (784) | 7.2 (45) |
|  | [n=4178] | [n=2022] | [n=2156] | [n=227] | [n=3328] | [n=623] |
| Smoking (ex), % (#) | 24.6 (1028) | 29.4 (594) | 20.1 (434) | 3.1 (7) | 25.0 (833) | 30.2 (188) |
|  | [n=4178] | [n=2022] | [n=2156] | [n=227] | [n=3328] | [n=623] |
| Num. cig daily<sup>2</sup> | 15.0 (10.0, 20.0) | 15.0 (10.0, 20.0) | 10.0 (8.0, 15.0) | 10.0 (1.0, 11.5) [n=14] | 15.0 (10.0, 20.0) | 11.0 (10.0, 18.8) |
| Med. (IQR) | [n=788] | [n=394] | [n=394] |  | [n=736] | [n=38] |
| Num. drinks daily<sup>3</sup> | 0.6 (0.2, 1.2) | 0.6 (0.2, 1.5) | 0.2 (0.0, 0.6) | 0.2 (0.1, 0.6) | 0.6 (0.2, 1.2) | 0.6 (0.2, 1.5) |
| Med. (IQR) | [n=3305] | [n=1723] | [n=1582] | [n=143] | [n=2749] | [n=413] |
| **Members household** |  |  |  |  |  |  |
| Living alone (1), % (#) | 12.7 (522) [n=4120] | 11.4 (228) [n=1994] | 13.8 (294) [n=2126] | 0.4 (1) [n=223] | 9.9 (328) [n=3302] | 32.3 (193) [n=598] |
| 2, % (#) | 34.9 (1438) [n=4120] | 34.8 (693) [n=1994] | 35.0 (745) [n=2126] | 2.7 (6) [n=223] | 34.0 (1123) [n=3302] | 51.7 (309) [n=598] |
| 3, % (#) | 18.1 (745) [n=4120] | 18.6 (371) [n=1994] | 17.6 (374) [n=2126] | 16.1 (36) [n=223] | 20.2 (668) [n=3302] | 6.9 (41) [n=598] |
| 4, % (#) | 20.8 (855) [n=4120] | 21.5 (428) [n=1994] | 20.1 (427) [n=2126] | 43.5 (97) [n=223] | 22.2 (734) [n=3302] | 4.0 (24) [n=598] |
| 5, % (#) | 8.1 (332) [n=4120] | 8.2 (163) [n=1994] | 7.9 (169) [n=2126] | 24.2 (54) [n=223] | 7.9 (262) [n=3302] | 2.7 (16) [n=598] |
| >5, % (#) | 5.5 (228) [n=4120] | 5.6 (111) [n=1994] | 5.5 (117) [n=2126] | 13.0 (29) [n=223] | 5.7 (187) [n=3302] | 2.5 (15) [n=598] |
| Senior care home<sup>4</sup>, % (#) | 0.3 (13) [n=4203] | 0.05 (1) [n=2032] | 0.6 (12) [n=2171] | 0.0 (0) [n=227] | 0.0 (0) [n=3343] | 1.9 (12) [n=633] |

<sup>1</sup>Education years were computed as number of years at school plus years at vocational schools and/or university including doctoral time, if applicable.
<sup>2</sup>Number of cigarettes smoked daily are provided for current smokers
<sup>3</sup>Number of alcoholic drinks for individuals drinking any alcohol (computed from the frequency of drinking and number of drinks when drinking, with a drink being equivalent to 0.33 l beer, 0.125 l wine, or 4 cl hard liquor)
<sup>4</sup>Residence in senior care homes has been not been assessed via questionnaire but from checking addresses of all study participants

### Self-reported positive PCR-based test for SARS-CoV-2

Of the 4 203 participants, 518 (12.32%) reported to have previously been tested by PCR for SARS-CoV-2 when filling out the questionnaire, including 74 reporting a positive PCR test result (1.76% of participants, **Table 2**). Of the 74 individuals with a reported previous test and positive PCR result, 66 were registered at the local health authorities with a positive PCR test resulting in a confirmed PCR positivity for 1.57% of the study participants. The remaining 8 individuals were not registered as being tested by local health authorities; these individuals may have misinterpreted the question, or were tested in a different county and/or site (airport, train station etc.).

**Table 2.** PCR-test self-report and confirmed positive PCR vs serostatus.

| **PCR test status** | **Serostatus negative**<br>**% (#)** | **Serostatus positive**<br>**% (#)** |
| --- | --- | --- |
| No test, n=3683 | 93.32 (3437) | 6.68 (246) |
| Test negative, n=427 | 89.00 (380) | 11.01 (47) |
| Test positive, n=74 | 6.76 (5) | 93.24 (69) |
| Test positive, confirmed, n=66* | 6.06 (4) | 93.94 (62) |
| Test status unknown, n=17 | 94.12 (16) | 5.88 (1) |
| Overall, n=4201 | 91.36 (3838) | 8.64 (363) |

\* 66 out of 74 self-reported PCR-test positive individuals were confirmed by local health authorities.

Among the 64 643 residents of the Tirschenreuth county aged ≥ 14years (numbers provided by local municipalities of the Tirschenreuth county), health authorities report 1.71% registered SARS-CoV-2 cases until June 2nd (n=1 108). This is similar to the observed fraction of study participants reporting a positive test result, which supports the notion that individuals with known infection were as likely to participate as those without known infection.

Seroprevalence for SARS-CoV-2 was assessed by two commercial (Roche Diagnostics and Shenzhen YHLO Biotech) and one in-house assay [13]. Roche’s Cobas Elecsys CLIA test detects nucleoprotein-(N)-directed complete Ig levels including IgG, the YHLO CLIA test used detects IgG antibodies to the N- and S-protein and the in-house ELISA quantifies IgGs binding to the receptor-binding domain (RBD) of the spike protein. Seropositivity was detected in 8.38% (95%-CI 7.85-9.26; n=4 187), 8.93% (95%-CI 8.11-9.14; n=4 186) or 9.17% (95%-CI 8.33-10.08; n=4 200) for the YHLO, the Roche-Cobas, and the in-house ELISA test, respectively (**Table 3**) among the individuals with a successful test result, respectively. Differences in the detected proportions might be attributed to (i) different specificities and sensitivities of the assays, (ii) different antigen-specific antibody levels, and/or (iii) different isotype distributions depending on the time since infection.

**Table 3.** Antibody status. Absolute (#) and relative (%) frequencies of individuals tested positive for SARS-CoV-2 antibodies using three different antibody tests accompanied by their 95% confidence interval (95%-CI)

| Test | ELISA<br>n=4200 | Roche-Cobas<br>n=4186 | YHLO<br>n=4187 |
| --- | --- | --- | --- |
| **Positive #**<br>**(%)** | 385<br>(9.17) | 374<br>(8.93) | 351<br>(8.38) |
| **95%-CI (%)<sup>1</sup>** | 8.33 – 10.08 | 8.11 – 9.84 | 7.85 – 9.26 |

<sup>1</sup>Confidence intervals (CI) were estimated according to Wilson-Score-Method.

### Latent Class Analysis to estimate true seropositives in TiKoCo study cohort

LCA to define seropositivity for all 4201 study participants with at least one result from any of the three antibody tests resulted in 363 individuals who scored SARS-CoV2 antibody positive (8.64%). This LCA-based analysis of the true antibody status on the basis of the three antibody tests was possible as the local independence assumption of the LCA holds (Garrett and Zeger’s log odds ratio test for a violation of the assumption yielded p-values for three pairwise comparisons of antibody tests of 0.98, 0.87 and 0.81, respectively). In addition, goodness of fit statistics of the model resulting from LCA indicated a high accuracy of the model (BIC=59.7 and CAIC=66.7) [17].

### Seroprevalence, dark figure factor and infection fatality ratio in the county Tirschenreuth

Seropositivity was 8.57% in men (n=174) and 8.71 % in women (n=189) of all tested study participants (n=4 201) **(Supplemental Table 2).** Within the various age groups of the study participants, seropositivity varied from 5.21% to 10.18% **(Supplemental table 3)**. Furthermore, the proportion of seropositive participants differed amongst the local municipalities of the county (**Supplemental Table 4**).

Taking into consideration the minor differences in the composition of our study sample compared to the Tirschenreuth population, the crude seroprevalences obtained for our study sample were standardized according to gender, age and local municipality to the county population, yielding a standardized overall seroprevalence of 8.57% (95%-CI 7.77-9.45), which was similar between women (8.64%; 95%-CI 7.54-9.89) and men (8.50%; 95%-CI 7.37-9.78) **(Figure 2a, Supplemental Table 2).**

**Figure 2.**
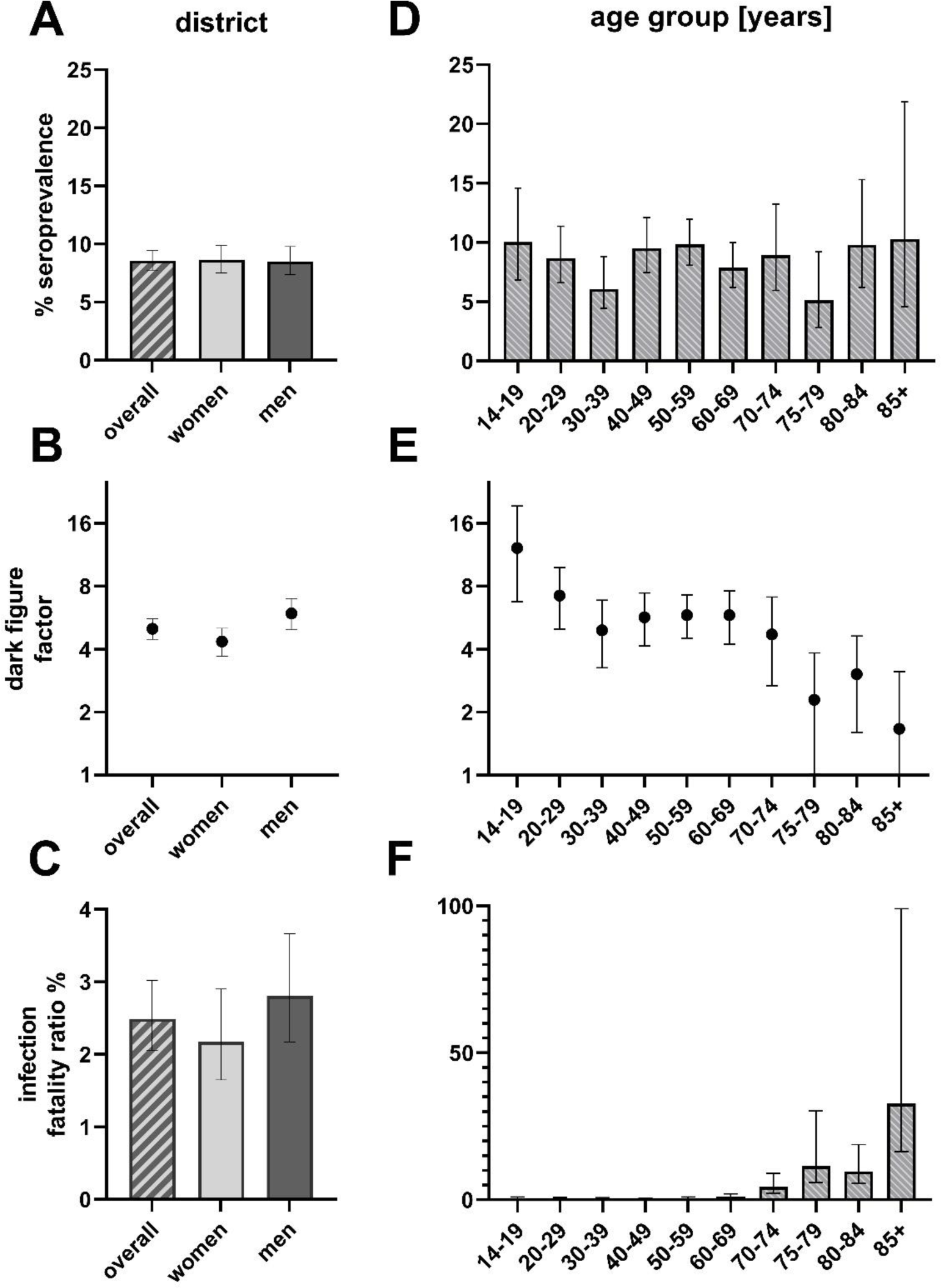
Standardized seroprevalence (%), dark figure factor and infection fatality ratio (%) in the overall county population (a-c) and the indicated age groups (d-f). Error bars represent 95% confidence intervals (95%-CI), respectively. *Figure was designed using GraphPad Prism version 8.4.3 for Windows, GraphPad Software, La Jolla California USA,* www.graphpad.com

By June 2nd, 2020, health authorities registered 1 108 PCR positive COVID-19 cases (1.71%) and 138 COVID- 19 related deaths in the Tirschenreuth population aged 14 years and older (64 643 inhabitants ≥14 years). Based on the standardized seroprevalence of 8.57%, 4 432 of 5 540 individuals calculated to be seropositive in the district of Tirschenreuth had not been registered as COVID-19 case based on a positive PCR test. Under the assumption that all ever infected produced antibodies detectable by our assessment, this indicated that 80.00% of infections had remained undetected by the massive PCR testing as performed in spring and early summer 2020 in this particular county **(Figure 2b, supplemental Table 2**). This corresponds to an underestimation of the cumulative number of infections by a factor of 5.00 in the ≥ 14-year-old population. Due to higher proportion of PCR+ women (1.98%) versus men (1.47%), the dark figure factor of undetected infections differed between women (factor 4.35) and men (factor 5.92).

Of note, of the 66 registered PCR-positive study participants (74 according to self-report), we found 4 (5 of 74) without antibodies (6.06% and 6.76%, respectively) (**Table 2**). This could be due to a false positive PCR test result, a primary failure to raise antibodies after infection, or a secondary loss of antibodies between seroconversion and blood draw end of June/early July, 2020.

In view of the 138 people who have died from or with COVID-19 in the county until June 2^nd^ (all aged ≥14 years), we calculated the IFR as 2.49% (95%-CI 2.06-3.02) for the population of the county Tirschenreuth aged 14 years and older, with an IFR of 2.18% (95%-CI 1.65-2.90) for women and 2.81% (95%-CI 2.17-3.67) for men (**Figure 2c, supplemental Table 2**). Of note, 62 of the 138 death cases in that county were senior citizens who lived in senior care homes (see below).

### Seroprevalence, dark figure and infection fatality ratio by age groups

We found gender- and municipality-standardized seroprevalences to vary from 5.14% (95%-CI 2.84-9.19) to 10.27% (95%-CI 4.56-21.87) by age groups (**Figure 2d, Supplemental Table 3**). When comparing the number of expected seropositives in the study population by age groups with the respective number of reported PCR positive individuals, the dark figure factor varied in the extremes from 12.15 (95%-CI 6.75-19.32) among teenagers (14-19 years) to 1.67 (95%-CI 1.00-3.12) in the elderly above 85 years, meaning that 91.75% and 40.12% of infections were unregistered in these two groups, respectively. For the other age groups, the age group specific dark figures varied between 3.04 to 7.21 **(Figure 2e, Supplemental Table 3**).

Based on the number of individuals estimated to be seropositive in the Tirschenreuth population by age group, we also determined the age-dependent IFR **(Figure 2f, Supplemental Table 3)**. With the exception of one person among the 20- to 29-year-old, no death related to COVID-19 was reported to health authorities for persons under 50 years of age. IFRs were low to moderate, <0.5% up to 59 years of age and 0.98% (95%- CI 0.49-2.04) for those aged 60 to 69 years, but then dramatically increased with age exceeding 10% (11.04%) for individuals older than 75 years (95%-CI 5.84-30.21) and 30% (31.55%, 95% CI 16.48-99.1%) for the elderly of 85 years and older. Such high IFRs may be attributed to co-morbidities in the older age groups, but might have been also aggravated by outbreaks seen in residential and nursing homes (see below).

### Seroprevalence by municipalities

In our study sample, seroprevalences differed markedly between the municipalities of the Tirschenreuth county (**Figure 3a)**. Being aware of sparse numbers of inhabitants for some smaller municipalities, we nevertheless took an effort to standardize our crude seroprevalence estimates to the study population by municipality, to better understand the regional infection dynamics in spring 2020. Standardized seroprevalences ranged from 22.65% (95%-CI 14.42-33.69) to 1.00% (95%-CI 0.16-5.47) (**Figure 3a, Supplemental Table 4)**. Interestingly, we observed a gradient in seroprevalences from eastern (high) to western (low) municipalities, separated by a forest belt. In addition to geographic barriers, patterns of municipalities with high seroprevalence and others with moderate to low seroprevalence may be explained by a wide range of aspects: founding effects by ski-travelers returning from Italy or Austria, the local character of beer festivities prior to and during carnival season, proximity of public PCR test centers and/or family doctors as well as growing awareness of infection occurrence followed by rigid testing.

**Figure 3.**
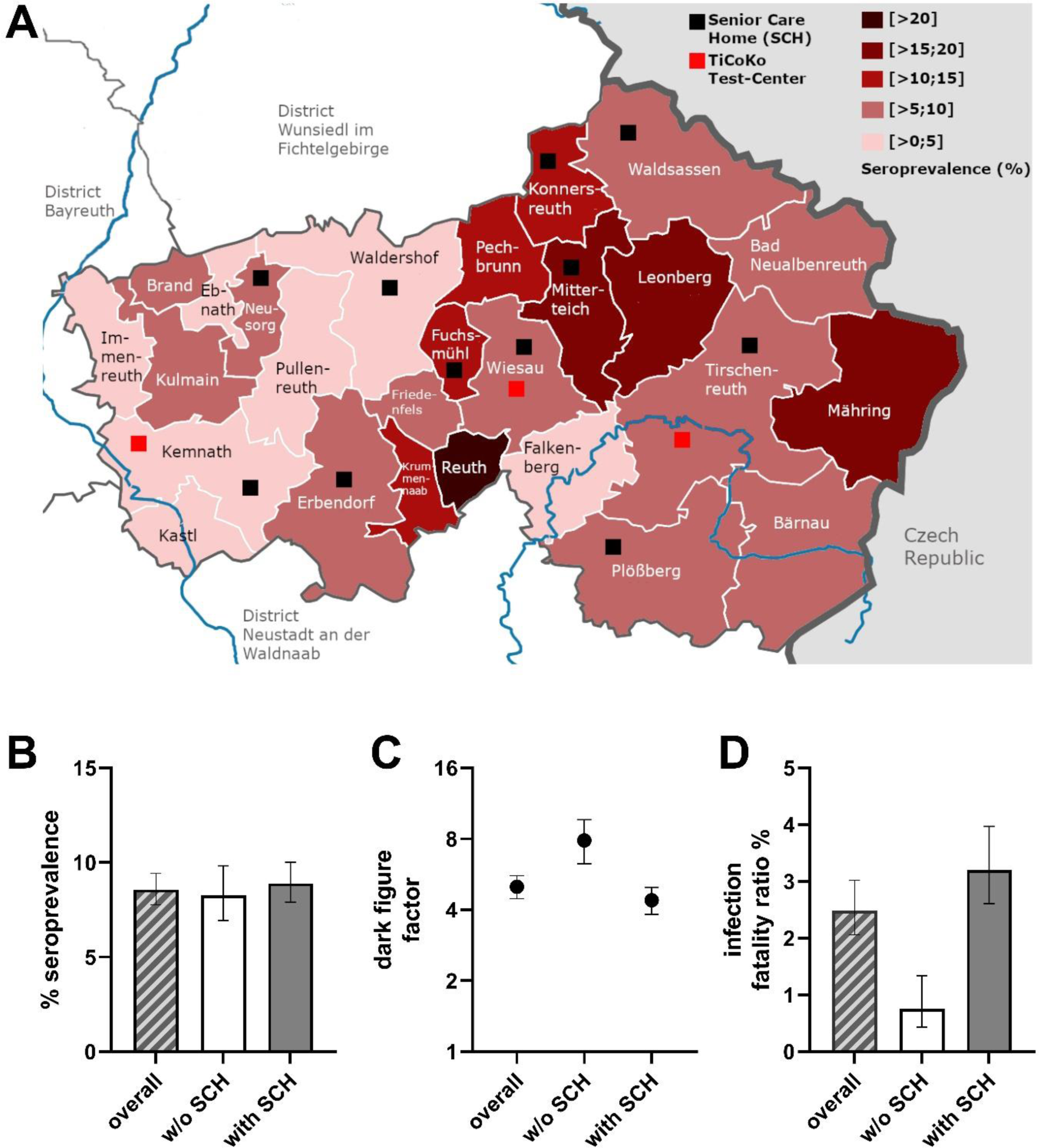
(a) Standardized seroprevalence (%) determined for inhabitants of the local municipalities in the county Tirschenreuth. (b-d) Standardized seroprevalence (b), dark figure factor (c) and infection fatality ratio (d) in the overall county population, in the population of local municipalities without senior care homes (w/o SCH) and with senior care homes (with SCH). 95% confidence intervals (95%- CI) are indicated, respectively. Red squares indicate the location of the study test centers, black squares highlight the municipality association of senior care homes. *Figure was designed using GraphPad Prism version 8.4.3 for Windows, GraphPad Software, La Jolla California USA,* www.graphpad.com*, and GIMP 2.10.22 for Windows, The GIMP Development Team, California USA,* www.gimp.org

Due to sparse numbers for some municipalities, we refrained from demonstrating dark figure factors and IFR for individual municipalities. An important question was the extent to which the 62 COVID-19 related deaths observed in senior care homes by June 2^nd^ compared to a total of 138 COVID-19 related death cases in the complete county may have contributed the IFR. Acknowledging previous reports on the high death toll in senior care homes, we separated municipalities into those with any senior care home and those without (11 versus 15, respectively) (**Figure 3a, Supplemental Table 5**). For this, we found an age- and sex-standardized seroprevalence of 8.90% (95%-CI 7.89-10.01) and 8.26% (95%-CI 6.93-9.83), respectively **(Figure 3b).** Considering the numbers registered PCR positive cases (888 versus 219). This translated into a dark figure factor of 4.08 (95%-CI 3.83-4.99) and 8.48 (95%-CI 6.28-9.66) for the municipalities with and without senior care homes, respectively **(Figure 3c).** This marked difference may – in part – be explained by extensive PCR testing in senior care homes including social contacts of employees and senior citizen residents. When calculating the expected number of seropositives in these two groups of municipalities (3904 and 1725, respectively) and comparing it to the number of deaths (125 and 13, respectively), we found IFRs of 3.20 (95%-CI 2.60-3.97) and 0.75% (95%-CI 0.44-1.33), respectively **(Figure 3d).** Thus, this underscores the pivotal role of senior care homes on population-based estimates of IFRs.

### Sensitivity analysis focusing on individuals not living in senior care homes

A large proportion of the observed COVID-19 related deaths were observed in senior care home (n=62 of the 124 deaths among individuals 70+ years of age, assuming senior care home residents are 70+). While individuals living in senior care homes (n=920, 1.42% of the population ≥ 14 years) were part of our random drawing of invited participants to capture the full population, this is a very specific group of individuals in terms of comorbidities and living situation. The computation of interpretable IFRs for individuals living in nursing homes alone by our design is challenged by low response (n=13, 12.15% of 107 invited residence inhabitants).

To understand the impact of senior care homes on IFRs, we computed sex- and municipality standardized seroprevalences and IFR estimates for individuals aged 70+ not living in senior care homes and compared these to the estimates for all individuals aged 70+ (combining age groups 70-74, 75-79, 80-84, 85+ from the previous analysis). We observed similar seroprevalence in this restricted analysis for individuals aged 70+ (7.38%, 95%-CI 5.71-9.57) compared to all 70+ (7.79%, 95%-CI 6.06-10.03). The IFR for individuals 70+ not living in senior care homes was 7.54% (95%-CI 5.34-11.28; #deaths=62), which was substantially lower than the IFR for those aged 70+ including senior care home residents (13.19%, 95%-CI 9.87-18.71, #deaths=124).

We also evaluated how the total IFR across all age groups was affected when omitting individuals living in senior care homes: again, there was no impact on standardized seroprevalence across all ages (8.47, 95%-CI 7.67-9.33; compared to 8.57%, 95%-CI 7.77-9.45, see above). The IFR for individuals across all ages omitting those living in senior care homes was 1.41% compared to the 2.49% in the total study population derived above.

### Reported previous illness

A “less healthy” elderly population in Tirschenreuth as compared to other areas in Germany could explain some of the excess of COVID-19 deaths observed. Thus, we compared participants of our study aged 70+ with a reference study, the population-based AugUR study participants from a geographically nearby region in Bavaria [12]. Participants of the AugUR study were residents from Regensburg city and Regensburg county and per design all aged 70+ (median age, 25^th^-75^th^ percentile 78.9, 75.7-82.5 years (AugUR) versus 77.0, 72.0- 81.0, this study). While the questions asked in our study here for self-reporting comorbidities were aligned to the questions answered by AugUR study participants, but AugUR had an in-person interview. For reported previous illnesses such as obesity, cardiovascular disease, type 2 diabetes, and lung disease, the respective proportions between the two studies showed no major differences (**Table 4**); reports of cancer, chronic kidney disease, and hypertension were more common in AugUR compared to TiKoCo participants aged 70+, which may be partly explained by the different mode of administering the questions. The overall picture in comparison to AugUR does not provide evidence for the TiKoCo participants to be more affected by previous illnesses reported as predisposing to severe COVID-19 [19].

**Table 4.** Previous illnesses reported by participants. Participants were asked whether they ever had a physician diagnosing any of the following diseases, except for obesity, which was derived from self-reported height and weight. Relative (%) and absolute (#) frequencies based on information of [n] participants.

|  | TiKoCo study participants |  |  |  |  |  | AugUR study participants<sup>a</sup> |
| --- | --- | --- | --- | --- | --- | --- | --- |
|  | All | Male | Female | Age 14-19 | Age 20-69 | Age ≥ 70 | Age ≥ 70 |
| Cancer, % (#) | 5.0 (204) [n=4100] | 4.6 (90) [n=1977] | 5.4 (114) [n=2123] | 0.0 (0) [n=222] | 3.7 (120) [n=3258] | 13.5 (84) [n=620] | 27.6 (672) [n=2439] |
| Kidney Dis., % (#) | 3.4 (138) [n=4100] | 3.7 (74) [n=1977] | 3.0 (64) [n=2123] | 0.5 (1) [n=222] | 2.7 (89) [n=3258] | 7.7 (48) [n=620] | 25.6 (624) [n=2439] |
| Obesity, % (#) | 27.0 (1122) [n=4154] | 28.0 (562) [n=2010] | 26.1 (560) [n=2144] | 7.5 (17) [n=226] | 27.6 (913) [n=3306] | 30.9 (192) [n=622] | 30.2 (734) [n=2432]<sup>b</sup> |
| Cardiovasc. Dis., % (#) | 9.9 (404) [n=4100] | 12.0 (238) [n=1977] | 7.8 (166) [n=2123] | 2.3 (5) [n=222] | 7.0 (228) [n=3258] | 27.6 (171) [n=620] | 28.0 (677) [n=2420] |
| Type-2 diabetes, % (#) | 7.7 (314) [n=4100] | 8.7 (172) [n=1977] | 6.7 (142) [n=2123] | 0.5 (1) [n=222] | 5.7 (185) [n=3258] | 20.6 (128) [n=620] | 21.4 (522) [n=2440] |
| Lung diseases, % (#) | 10.5 (432) [n=4100] | 10.6 (209) [n=1977] | 10.5 (223) [n=2123] | 9.5 (21) [n=222] | 10.0 (325) [n=3258] | 13.9 (86) [n=620] | 12.4 (302) [n=2434]<sup>c</sup> |
| Hypertension, % (#) | 29.9 (1227) [n=4100] | 33.3 (659) [n=1977] | 26.8 (568) [n=2123] | 1.4 (3) [n=222] | 26.1 (850) [n=3258] | 60.3 (374) [n=620] | 70.6 (1723) [n=2440] |
| Blood clotting, % (#) | 2.2 (91) [n=4100] | 2.0 (39) [n=1977] | 2.4 (52) [n=2123] | 0.9 (2) [n=222] | 1.6 (52) [n=3258] | 6.0 (37) [n=620] | n.a. |
| Autoimmune dis., % (#) | 7.1 (291) [n=4100] | 4.1 (81) [n=1977] | 9.9 (210) [n=2123] | 1.4 (3) [n=222] | 7.1 (232) [n=3258] | 9.0 (56) [n=620] | n.a. |

<sup>a</sup> AugUR (self-reported information at last visit): age 80.04 +/- 5.11 (median 79.39, range 70.4 – 98.3); 52.1% female
<sup>b</sup> measured at study center; BMI>30 kg/m<sup>2</sup>
<sup>c</sup> chronic bronchitis or asthma

### Reported symptoms and hospitalization

Among the 4 162 individuals providing information on the experience of any COVID-19 related symptoms since the start of the pandemic (as per Feb 1st, 2020), 3799 were negative for SARS-CoV-2 specific antibodies and 363 were seropositive (**Figure 4a**). Interestingly, 12.9% (n=47) of the seropositive individuals reported no symptom at all (i.e. asymptomatic infected), 82.4% (n=299) reported at least one symptom, but did not report hospitalization due to COVID-19, and 4.7% (n=17) reported hospitalization due to severe COVID-19. We found almost all symptoms more frequently reported among seropositive subjects compared to seronegatives, except eye inflammation and headache (highest Odds Ratios, OR, for olfactory or taste problems and fever, OR=21.94, 16.18, or 6.54, respectively) (**Figure 4b; Supplemental Table 6**). These results support a role of most of these symptoms for SARS-CoV-2 infection, with a predominant role of olfactory and taste impairment. Reported symptoms in all participants indicate small differences by sex and age groups (**Supplemental table 7**).

**Figure 4.**
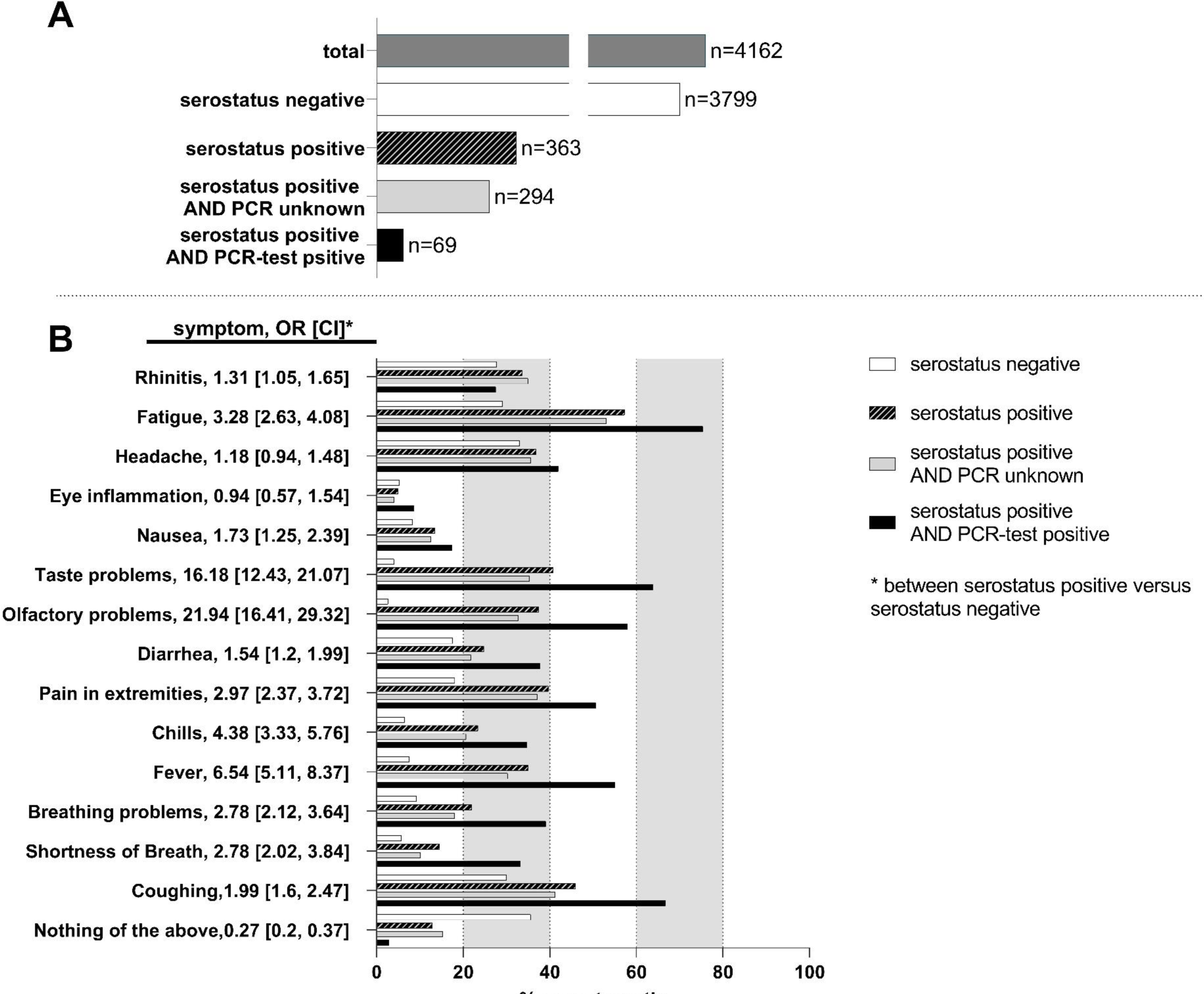
(a) Number of individuals responding to the question regarding symptoms (total), categorized into individuals with either negative serostatus or individuals determined serostatus positive including the indicated subgroups. (b) Percent individuals of the indicated categories, who developed one or several of the indicated symptoms. Odds Ratios, OR, as well as 95%-CI [CI] testing serostatus positive versus serostatus negative (unadjusted) are depicted. Results adjusted for age and gender are very similar (data not shown). *Figure was designed using GraphPad Prism version 8.4.3 for Windows, GraphPad Software, La Jolla California USA,* www.graphpad.com

These observations persisted when focusing on participants with positive antibody status, who did not report a positive SARS-CoV-2 PCR test results (not tested or tested negative, i.e. unaware infected), though most symptoms were reported less frequently than among participants with positive antibody status and report of positive SARS-CoV-2 test (i.e. known infected, **Supplemental Table 6**). One exception was the report of rhinitis, which was more frequent among the unaware infected than among the known infected and substantially more than among the antibody-negatives. This would be in line with an underestimation of rhinitis in its role of SARS-CoV-2 infection.

Interestingly, symptoms were experienced clearly more frequently not only among the known infected, but also among the unaware infected when compared to the antibody-negatives **(Figure 4b).** The fact that the unaware infected report symptoms less frequently than known infected is in line with the fact that asymptomatic or mild symptomatic infected were missed in the targeted testing (at least at this early stage of the pandemic in spring and early summer 2020).

We found also a strong association between individuals reporting a severe bronchitis/pneumonia (i.e. needing bed rest or physician or hospitalization) since the start of the pandemic (as per Feb 1^st^, 2020): the

ORs for being antibody-positive among those reporting bronchitis/pneumonia related bed-rest or physician or hospitalization were 3.4, 3.2, or 15.4, respectively (**Table 5**). Most of the antibody-positive individuals with bronchitis/pneumonia requiring hospitalization were aware of their infection (n=11 among the 14 with hospitalization and antibody-positive). The majority of individuals reporting hospitalization for bronchitis/pneumonia since the start of the pandemic was infected (14 of the 24).

**Table 5.** Report of bronchitis/pneumonia by serostatus. Participants were asked whether they had experienced a bronchitis or pneumonia since the start of the pandemic (as per Feb 1^st^, 2020) and to what degree they had been affected. Bronchitis/pneumonia are the lead diseases with which individuals are hospitalized that are potential patients of COVID-19. We present the relative (%) and absolute (#) frequencies of individuals with Bronchitis in the overall sample and for different combinations of seropositivity and self-reported PCR test results and quantify the association of seropositivity with Bronchitis based on Odds ratios between seropositive and seronegative individuals (OR, with associated 95%-confidence interval, CI).

| **Bronchitis (since Feb)** | **Overall** | **Seropositive AND PCR-test pos.** | **Seropositive AND no PCR-test or neg.** | **Seropositive** | **Seronegative** | **OR [95%-CI]** |
| --- | --- | --- | --- | --- | --- | --- |
| No % (#) | 91.3 (3799) [n=4162] | 70.1 (47) [n=67] | 83.2 (243) [n=292] | 80.8 (290) [n=359] | 92.3 (3509) [n=3803] | 0.35 [0.26, 0.47] |
| Affected, a little % (#) | 4.4 (182) [n=4162] | 4.5 (3) [n=67] | 5.5 (16) [n=292] | 5.3 (19) [n=359] | 4.3 (163) [n=3803] | 1.25 [0.77, 2.03] |
| Affected, stayed in bed % (#) | 1.5 (63) [n=4162] | 4.5 (3) [n=67] | 4.1 (12) [n=292] | 4.2 (15) [n=359] | 1.3 (48) [n=3803] | 3.41 [1.89, 6.15] |
| Affected, needed physician % (#) | 2.3 (94) [n=4162] | 4.5 (3) [n=67] | 6.2 (18) [n=292] | 5.8 (21) [n=359] | 1.9 (73) [n=3803] | 3.17 [1.93, 5.22] |
| Affected, hospitalized % (#) | 0.6 (24) [n=4162] | 16.4 (11*) [n=67] | 1.0 (3) [n=292] | 3.9 (14) [n=359] | 0.3 (10) [n=3803] | 15.39 [6.79, 34.91] |

Among the 18 individuals reporting to have been hospitalized since Feb 1<sup>st</sup>, 2020, due to COVID-19 disease (and having had a positive PCR-test), 11 reported to have been hospitalized due to bronchitis or pneumonia.

### Association of demographic and lifestyle factors with seropositivity

We were interested in the question whether demographic characteristics were associated with higher seropositivity and thus higher probability of SARS-CoV-2 infection. We evaluated the association of age, sex, education years, size of household, or having worked in a profession with higher probability of contact (during the spring 2020 lockdown in Tirschenreuth, medical/nursing profession and cashiers/salesperson in groceries) with seropositivity in a logistic regression model. We found no evidence for association with age, sex, education or household size (P-values > 0.05) (**Figure 5, Supplemental Table 8**). With regard to occupation, we found no increased seropositivity among cashiers/salespersons in groceries, but a doubling of the odds of being antibody-positive among medical and nursing professions compared to other occupations (OR=2.26, 95%-CI: 1.53-3.28, **Figure 5**).

**Figure 5.**
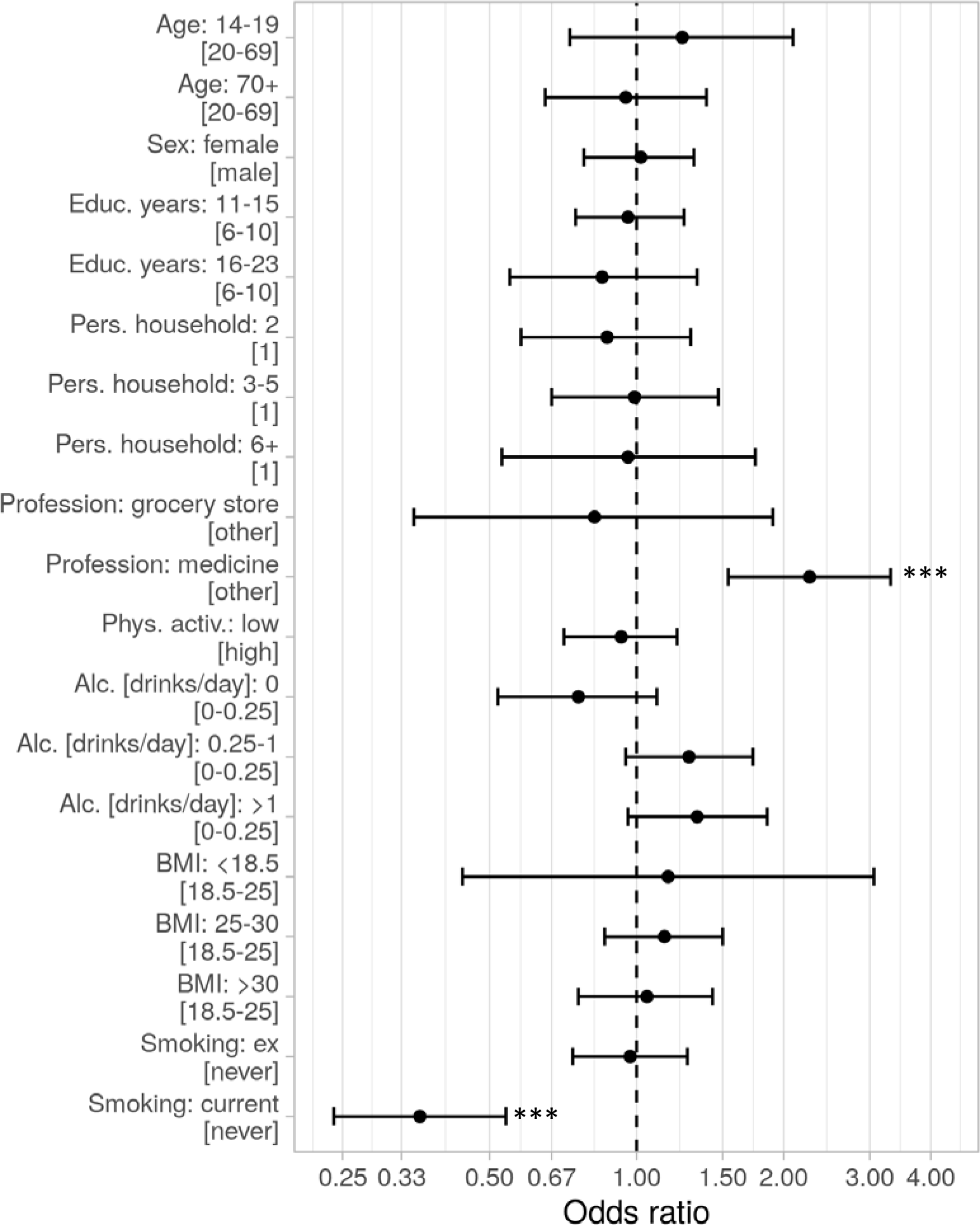
Association of demographic and life style factors with seropositivity. Odds ratios including the 95%-CI are indicated, respectively. P < 0,001 (***). *Analysis was conducted in R (R Core Team (2020). R: A language and environment for statistical computing. R Foundation for Statistical Computing, Vienna, Austria. URL* http://www.R-project.org/*). This figure was produced using the package ggplot2 (Wickham, H. (2009) ggplot2: elegant graphics for data analysis. Springer New York)*.

We were also interested whether seropositivity was associated with factors capturing a less healthy lifestyle (low physical activity, current or ex-smoking, drinking alcohol, or increased body-mass-index as a marker for excess calorie intake). We found no association with any lifestyle factor, except for current smoking. Remarkably, the odds of being seropositive was substantially decreased for current smokers compared to never-smokers (OR=0.36, 95%-CI: 0.24-0.53), while there was no association for ex-smokers (**Supplemental Table 8**). This would correspond to a 2.8-fold increase in the odds of being seropositive among never-smokers or ex-smokers compared to current smokers **(Figure 5).**

### Sensitivity analyses for the detected strong association of current smoking as being protective against being seropositive

The finding that current smoking was associated with a smaller probability of being seropositive is counter- intuitive under a hypothesis that smoking was associated with a less healthy attitude and more risky behavior. We conducted several sensitivity analyses in the search for potential confounding effects: first, we found stable associations across age groups and sex (unadjusted OR of seropositivity for current smokers vs. ex-/never smokers = 0.43, 0.34, and 0.47 for age groups 14-39 (young), 40-59 (mid), 60+ (old), respectively; 0.32 for men and 0.51 for women) (**Supplemental Table 9**). Second, we considered the possibility that the association of current smoking with reduced probability of seropositivity was due to an impaired ability of smokers to raise antibodies based on a previous report[20,21]. However, among the 74 individuals reporting a positive SARS-CoV-2 PCR test, we observed only 5 individuals without antibodies and these included 1 current smoker and 4 never-smokers (20% smoker as in the full sample). Furthermore, the association persisted when restricting to individuals having reported a previous PCR test: current smoking was associated with a positive test report versus negative test report (among the n=501 tested: OR=0.35), which is in line with a hypothesis that current smoking was associated with a lower risk of infection (**Supplemental Table 10)**. This finding was not compromised by a higher proportion of current smokers among those tested, as previously reported by others [22]: 21.0% of those tested and 20.2% of those not tested were smokers (OR=1.07 adjusted for age, sex). Of note, we observed more women than men tested (14.4% and 9.4%, respectively), but no difference across age groups (9.7%, 12.2%, 11.5%, for age 14-19, 20-69, 70+, respectively). Finally, we also found a significant dose-response between the number of daily smoked cigarettes and seropositivity among all participants (zero cigarettes for never and ex-smokers OR=0.50 per 10 cigs/day, 95%-CI: 0.37-0.65, modelling reported # cigarettes smoked daily in a linear fashion, adjusted for age and sex) and a similar association restricting to current smokers (OR=0.69 per 10cigs/day, 95%-CI: 0.43- 1.07, **Supplemental Table 11**). Allowing for non-linear dose-response supported this finding (**Supplemental Figure 2)**.

A reverse epidemiology effect could induce a bias here, when individuals being current smokers at the height of the outbreak included severely ill individuals (e.g. cancer, severe heart disease) that prevented individuals from going outside or primed individuals to be particularly careful avoiding infection. However, the smoking-associated severe diseases would affect more likely older individuals and are less likely among younger individuals; therefore, the stable effect estimates across age groups provide evidence against such bias that would fully explain the strong association across age groups. Another potential bias needs to be considered when smokers who were infected were less likely to participate in the study. However, most individuals have not known their seropositivity at the time of questionnaire completion and participation.

## Discussion

Data on the seroprevalence for SARS-CoV-2 infections from population-based studies is superior to convenience sample data, but such data is limited in Germany. Our population-based study recruited 4 203 individuals aged at least 14 years from the county hit the most and very early by the pandemic in Germany in spring 2020. We had invited a random sample of Tirschenreuth county inhabitants and achieved a high response at 64%. We derived a standardized sero-prevalence of SARS-CoV-2 of 8.6% with little variation across age groups, lower among current smokers and higher among individuals working in medical or nursing profession. Age-specific IFRs were < 0.5% for age groups below 60 years, 1.0% for those aged 60-69, 13.2% for those aged 70+ and overall IFR across age groups was 2.5%. We found a substantial impact by COVID-19 related deaths in senior care home residences, which comprised almost 46% of Tirschenreuth county deaths, reflected by an IFR of 7.5% among individuals aged 70+ and an overall IFR of 1.4% when excluding senior care home residence from the computation.

The seroprevalence of 8.5% implied that only one out of five SARS-CoV-2 infections had been registered by health authorities by community targeted testing at that time. While the seroprevalence and thus the implicated cumulative prevalence of infection was stable across all studied age groups, the lower number of registered infections in the young yielded a dark figure factor as high as 12.1 in 14 to 19-year-old compared to 1.7 in the ≥ 85-year-old. This is consistent with asymptomatic forms of COVID-19 at younger age and a testing strategy focusing on symptomatic individuals, particularly at this early phase of the pandemic in spring 2020. Our findings underscore the need for SARS-CoV-2 testing particularly in the young and suggests a note of caution to deduce infection rates from registered COVID-19 cases by a dark figure factor that is not adjusted for age.

We found several interesting aspects with regards to symptoms, in line with previous studies (e.g. [11,12,23]), particularly when comparing individuals unaware of their infection to the aware. We found a doubling of seroprevalence among individuals having worked in medical or nursing profession during the outbreak compared to other occupations, consistent to previous studies (e.g. [23]), which might reflect limited access to appropriate protective gear at that very early stage. We did not find higher seroprevalence in individuals working in groceries. Particularly important to discuss is the finding that current smoking compared to never smoking, but not ex-smoking, was associated with a decreased risk of being seropositive and for reported infection. The association is strong, stable across age groups and sex, and exhibited a dose-response-effect. While the sample size of smokers among individuals with reported PCR-test was limited, the stable risk estimates and the few individuals with reported infection not showing antibodies suggests that the finding was not due to a lack of antibody building, but due to lower infection probability. This does not allow for the conclusion that smoking per se was protective. While smoking is generally linked to a less healthy and more risky behavior, a behavior associated with smoking that guards against infection could result in the same observation, e.g. gathering socially more outside or less frequently. A lower seroprevalence among smokers was also found by other population-based studies[23,24]. A biological explanation for a protective effect of nicotine has been reported previously as the smoking paradoxon and a nicotine-receptor-related hypothesis[25,26]. This was based on the observation of a lower prevalence of smokers among individuals hospitalized due to COVID-19. However, individuals affected with severe COVID-19 requiring hospitalization are predominantly older individuals and there are fewer smokers among older individuals (7.2% in our study among those aged 70+, versus 23.6% among those aged 20-69) rendering age as a confounder in these analyses. Further functional studies are warranted to understand the reason for this observed lower seroprevalence and the lower PCR-test reported infections among smokers. Any conclusion that smoking was preventive for SARS-Cov-2 infection needs to be rigorously challenged and, if substantiated, conclusions made need to weigh in the adverse public health impact due to the severe other implications of smoking like lung cancer.

Our study was unique by employing three different test systems, two commercial assay platforms and one in-house assay, to determine SARS-CoV-2 specific antibody responses. These assay systems varied (i) regarding the viral target for the specific antibodies (S-RBD for the in-house assay, S and N protein for the YHLO assay and N protein only for the Roche assay), (ii) regarding the technical setup (*in-house* ELISA versus 2 commercial CLIAs) and (iii) regarding the detected antibody Isotypes (IgG versus total Ig including IgA and IgM). Furthermore, all 3 assay systems varied according to the manufacturers specifications in terms of clinical sensitivity and specificity. Not unexpectedly, the crude seropositivity varied among the three platforms from 8.38 % to 9.17 % in the study participants. In order to determine the true seropositivity, we employed LCA, a statistical approach that has repeatedly been used in the past to resolve discrepancies between diagnostic tests in the absence of an established gold standard [15]. The statistical model resulting from LCA shows an excellent fit to the observed pattern of results from the three antibody tests and fulfills the crucial assumption of local independence necessary to draw valid conclusions from the model. The model-derived definition of seropositivity uses the available information from all 3 antibody tests, combines it in an objective manner, and constitutes the basis for our computation of seroprevalences and associated measures like dark figure and IFR.

Based on registered COVID-19 related deaths obtained from health authorities and the number of Tirschenreuth inhabitants with infection during spring 2020 projected from standardized seroprevalence, we derived IFR estimates. As expected, IFR estimates substantially depended on age, resulting in nearly zero up to age 59 and 1.0%, 4.2%, 11.0%, 9.3%, and 31.5% for our age groups 60-69, 70-74, 75-79, 80-84, or 85+, respectively. While it is difficult to compare age-specific IFRs across published studies due to different age ranges included and different age groups reported, a recent meta-analysis derived a meta-regression model using age-specific IFRs across 27 population-based studies worldwide [27]. Based on their model, predicted IFR were < 0.5% for ages below 60 and 1.4%, 3.4%, 6.1%, 11.3%, or 21.9% at age 65, 72.5, 77.5, 82.5, and 88 years, respectively, reflecting the midpoints of our age groups. This is in line with our findings taking the confidence intervals into account.

The overall IFR at 2.49% was high compared to the reported IFR of 0.35% for Gangelt, another German hot spot early during the pandemic [7]. The difference in IFR may be, at least in part, explained by a smaller proportion of elderly inhabitants in the municipality Gangelt compared to Tirschenreuth county (9.5% versus 12.0% aged 75+)[28,29]. Our data suggests that another important determinant for the extent of IFR, besides pure age, is the inclusion of senior home residents. While 13.7% of the registered COVID-19 infections in Tirschenreuth and 46% of COVID-19 related deaths [9] were residents of senior care homes, only 7.6% of the registered COVID-19 cases were senior care home residents in Gangelt[7,9]. When separating the Tirschenreuth communities into those with and those without any senior care homes, we found a total IFR of 3.2% and 0.8%, respectively. Importantly, the IFR for individuals aged 70+ at 13.2% reduced to 7.5% when excluding senior care home residents from the deaths and the sero-positives. The deaths among senior care home residents also impacts the total IFR across all age groups: when excluding senior care home deaths and respective sero-positives, the total IFR is 1.4% rather than 2.5% originally. This underscores the dominant impact of senior care home residents in the computation of IFRs.

Concern has been raised for population-based studies that include senior care home residents to underestimate seroprevalence due to low response and thus to overestimate COVID-19 IFRs [10]. However, excluding senior care home residents clearly underestimates total IFR. Recruiting elderly, particularly senior care home residents, is a substantial challenge. For this reason, many population-based seroprevalence studies excluded individuals above the age of 70 years [30] or exclude senior care home residents by design (e.g.[31,32]) or by analysis [23]. Nevertheless, senior care home residents are a part of our populations. In our study, we included senior care home residents into our random sampling and provided mobile study teams to visit seniors in private home as well as senior care homes when appropriate. By this, we limited non-response in the elderly, but still observed a lower response compared to all.

It was hypothesized that the high Tirschenreuth death toll was potentially, at least in part, related to more frequent comorbidities or a less healthy lifestyle in this population[8,9]. However, self-reported lifestyle factors and comorbidities among Tirschenreuth study participants aged 70+, i.e. the age group where most COVID-19 related deaths occurred, compared well with self-report from individuals 70+ from another population-based study in a city in the vicinity of Tirschenreuth, in Regensbug [12]: study participants reported comparably frequently the pre-existence of type 2 diabetes, cardiovascular disease and had similar proportion of obesity. The only observation where participants differed was a higher proportion of individuals aged 70+ who were current smokers in Tirschenreuth compared to Regensburg and the number of cigarettes smoked per day was also higher on average. However, it is not plausible that this can explain the high death toll. The majority of deaths the hard-hit European countries have happened at that time in nursing homes [33] and a large proportion of deaths in the US also followed this pattern [34]. As suggested by our data, the high death toll in the county of Tirschenreuth and the relatively high IFR can be, at least in part, be attributed to outbreaks in senior care homes, particularly at this early stage of the pandemic in Europe. At the time of the outbreak, awareness of the extent of the problem as well as protective gear like masks for the community had been limited. Still, outbreaks in senior care homes continue to happen with devastating outcome, but will hopefully be controlled by targeted vaccination efforts.

The scheduled follow-up investigations of study participants will allow for further analyses directed towards monitoring the temporal development of these SARS-CoV-2 related metrics such as seroprevalence, dark figure factor and IFR in the study population as well as the durability of the antibody response to the SARS-CoV-2 infection. Our present results provide important and comprehensive information from a population-based epidemiological perspective that help to understand the COVID-19 pandemic.

## Author contributions

The study was conceptualized and supervised by RW and KÜ. KÜ, RW, IH and OG designed the study, ethics approval was granted to RW and KÜ. OG and KÜ conceptualized random sampling of study participants. IH, KS, RW, MK designed the self-reporting questionnaire. RW, MK, MB developed the database concept. MB, SB, HHN, AK, RW, KÜ, and HS coordinated the study centers and logistics.

Experimental data were generated, analyzed and interpreted by DP, AS, SE, RB, MT, CM, PS, KÜ and RW, data curation OG and RW. Statistical analysis was performed by OG, AP, FG, IH and HK. Study infrastructure was supported by AG, KÜ, HS and RW. RW, KÜ, FG, IH and OG wrote the manuscript, all co-authors contributed to writing, reviewed and approved the manuscript.

## Supporting information

Manuscript Tirschenreuth Supplemental Tables and Figures

## Data Availability

All authors declare that data and materials and codes will be made available.

## Acknowledgement

We are particularly grateful to all the study participants. We would like to thank the office staff and numerus students of the University of Regensburg and the University Erlangen as well as the employees of the Bavarian Red Cross, the members of the civil protection, the county office and the public health office of the county Tirschenreuth, respectively, for their tremendous support. We are also grateful to Christena Wolff from wECARE and Jakob Niggel from MaganaMed for providing Questionnaires and the database for the study.

## Availability of data and material (data transparency)

All authors declare that data and materials will be made available according to the guidelines of the journal.

## Code availability (software application or custom code)

Codes developed to generate and interpret the data will be made available.

## Consent to participate (include appropriate statements)

Informed consent was obtained from all individual participants included in the study.

## Consent for publication (include appropriate statements)

The authors affirm that human research participants provided informed consent for publication of the data presented in this study.

