## Supplementary material for "Estimates and determinants of SARS-CoV-2 seroprevalence and infection fatality ratio using latent class analysis: the population-based Tirschenreuth study in the hardest-hit German county in spring 2020": Manuscript Tirschenreuth Supplemental Tables and Figures

**Supplemental Table 1:** Distribution of the inhabitants of the county of Tirschenreuth and of the study participants (n = 64 643 of inhabitants of Tirschenreuth county aged ≥14 years and 4 203 study participants, respectively).

| | Number of inhabitants in the county | % (among county inhabitants aged ≥ 14 years) | Number of study participants | % (among study participants) | absolute deviation between study and county (%) |
|---|---|---|---|---|---|
| **Gender analysis** | | | | | |
| men | 32239 | 49.87 | 2032 | 48.35 | -1.52 |
| women | 32404 | 50.13 | 2171 | 51.65 | 1.52 |
| **Age group analysis** | | | | | |
| 14 - 19 | 3994 | 6.17 | 227 | 5.40 | -0.77 |
| 20 - 29 | 8146 | 12.58 | 523 | 12.44 | -0.14 |
| 30 - 39 | 8430 | 13.18 | 585 | 13.92 | 0.74 |
| 40 - 49 | 8782 | 13.56 | 601 | 14.30 | 0.74 |
| 50 - 59 | 12813 | 19.79 | 882 | 20.99 | 1.20 |
| 60 - 69 | 10412 | 16.08 | 752 | 17.89 | 1.81 |
| 70 - 74 | 3422 | 5.29 | 232 | 5.52 | 0.23 |
| 75 - 79 | 3350 | 5.17 | 192 | 4.57 | -0.60 |
| 80 - 84 | 3412 | 5.27 | 161 | 3.83 | -1.44 |
| ≥ 85 | 1882 | 2.91 | 48 | 1.14 | -1.77 |
| **Municipality analysis** | | | | | |
| Bad Neualbenreuth | 1186 | 1.83 | 93 | 2.21 | 0.38 |
| Bärnau | 2795 | 4.32 | 189 | 4.50 | 0.18 |
| Brand | 1025 | 1.58 | 59 | 1.40 | -0.18 |
| Ebnath | 1710 | 2.65 | 65 | 1.55 | -1.10 |
| Erbendorf | 4476 | 6.91 | 263 | 6.26 | -0.65 |
| Falkenberg | 822 | 1.27 | 73 | 1.74 | 0.47 |
| Friedenfels | 1103 | 1.70 | 87 | 2.07 | 0.37 |
| Fuchsmühl | 1387 | 2.14 | 87 | 2.07 | -0.07 |
| Immenreuth | 1600 | 2.47 | 100 | 2.38 | -0.09 |
| Kastl | 1208 | 1.87 | 83 | 1.97 | 0.10 |
| Kemnath | 4773 | 7.37 | 294 | 7.00 | -0.37 |
| Konnersreuth | 1501 | 2.31 | 109 | 2.59 | 0.28 |
| Krummennaab | 1299 | 2.01 | 85 | 2.02 | 0.01 |
| Kulmain | 1928 | 2.98 | 136 | 3.24 | 0.26 |
| Leonberg | 870 | 1.34 | 69 | 1.64 | 0.30 |
| Mähring | 1560 | 2.41 | 107 | 2.55 | 0.14 |
| Mitterteich | 5899 | 9.11 | 376 | 8.95 | -0.16 |
| Neusorg | 1810 | 2.80 | 131 | 3.12 | 0.32 |
| Pechbrunn | 1173 | 1.81 | 72 | 1.71 | -0.10 |
| Plößberg | 2835 | 4.38 | 206 | 4.90 | 0.52 |
| Pullenreuth | 1500 | 2.32 | 110 | 2.62 | 0.30 |
| Reuth b.Erbendorf | 994 | 1.54 | 71 | 1.69 | 0.15 |
| Tirschenreuth | 7807 | 12.06 | 547 | 13.01 | 0.95 |
| Waldershof | 3869 | 5.98 | 205 | 4.88 | -1.10 |
| Waldsassen | 5862 | 9.05 | 356 | 8.47 | -0.58 |
| Wiesau | 3651 | 5.64 | 230 | 5.47 | -0.17 |

**Supplemental Table 2.** Seroprevalence (SP), dark figure (DF) factor and infection fatality ratio (IFR) in the study population and standardized to the population of the county Tirschenreuth

| Group | SP (%) | SP (%) 95%-CI[6] | PCR +[2] (%) | DF[3] factor | DF[3] factor 95%-CI[7] | IFR[4] (%) | IFR[4] (%) 95%-CI[7] |
|---|---|---|---|---|---|---|---|
| **overall** | | | | | | | |
| crude | 8.641 | 7.828 - 9.529 | | | | | |
| standardized [1] | 8.571 | 7.768 - 9.449 | 1.710 | 5.013 | 4.462 - 5.587 | 2.487 | 2.058 - 3.022 |
| **men** | | | | | | | |
| crude | 8.567 | 7.427 - 9.864 | | | | | |
| standardized [2] | 8.500 | 7.373 - 9.783 | 1.434 | 5.928 | 4.978 - 6.939 | 2.806 | 2.171 - 3.669 |
| **women** | | | | | | | |
| crude | 8.710 | 7.595 - 9.970 | | | | | |
| standardized [2] | 8.641 | 7.540 - 9.888 | 1.984 | 4.355 | 3.707 - 5.038 | 2.175 | 1.648 - 2.903 |

[1] Standardized according to age, gender and municipality
[2] Standardized according to age and municipality
[3] Registered PCR positive (%) by local health authorities
[4] Dark figure factor: Ratio of standardized seroprevalence (%) and registered positive PCR (%), respectively.
[5] IFR (%): Infection fatality ratio. Percentage of people who have died from or with CoV-2 infection relative to the calculated number of seropositive individuals
[6]Confidence intervals (CI) were estimated according to Wilson-Score-Method
[7]Confidence intervals (CI) were computed as Bayesian credibility intervals (see methods)

**Supplemental Table 3.** Age-specific seroprevalence (SP), dark figure (DF) factor and infection fatality ratio (IFR) in the study cohort and standardized according to the gender and local municipality distribution of the population of the county Tirschenreuth

| Age group | SP (%)[1] | SP(%)[1]; 95%-CI[5] | SP (%)[2] | SP(%)[2]; 95%-CI[5] | Registered positive PCR (# and (%)) | DF[3] factor | DF[3] factor; 95%-CI[6] | Number of deaths | IFR[4] (%) | IFR[4] (%); 95%-CI[6] |
|---|---|---|---|---|---|---|---|---|---|---|
| **14-19** | 10.177 | 6.878 - 14.808 | 10.039 | 6.818 - 14.595 | 33 (0.826) | 12.154 | 6.749 - 19.322 | 0 | 0 | 0.000 - 0.993 |
| **20-29** | 8.795 | 6.659 - 11.533 | 8.676 | 6.588 - 11.367 | 98 (1.203) | 7.212 | 4.975 - 9.811 | 1 | 0.141 | 0.034 - 0.823 |
| **30-39** | 6.154 | 4.478 - 8.402 | 6.070 | 4.433 - 8.828 | 105 (1.231) | 4.931 | 3.258 - 6.845 | 0 | 0 | 0.000 - 0.749 |
| **40-49** | 9.651 | 7.540 - 12.274 | 9.519 | 7.455 - 12.098 | 147 (1.674) | 5.687 | 4.147 - 7.424 | 0 | 0 | 0.000 - 0.452 |
| **50-59** | 9.989 | 8.179 - 12.146 | 9.853 | 8.083 - 11.972 | 217 (1.694) | 5.818 | 4.522 - 7.247 | 5 | 0.396 | 0.172 - 0.956 |
| **60-69** | 7.979 | 6.249 - 10.136 | 7.871 | 6.179 - 9.990 | 141 (1.354) | 5.812 | 4.244 - 7.593 | 8 | 0.976 | 0.489 - 2.038 |
| **70-74** | 9.052 | 5.997 - 13.441 | 8.929 | 5.948 - 13.248 | 65 (1.899) | 4.701 | 2.678 - 7.089 | 13 | 4.255 | 2.289 - 8.961 |
| **75-79** | 5.208 | 2.853 - 9.321 | 5.138 | 2.847 - 9.189 | 75 (2.239) | 2.295 | 1.016 - 3.837 | 19 | 11.040 | 5.836 - 30.210 |
| **80-84** | 9.938 | 6.210 - 15.533 | 9.803 | 6.168 - 15.312 | 110 (3.224) | 3.041 | 1.605 - 4.622 | 31 | 9.268 | 5.563 - 18.820 |
| **85+** | 10.417 | 4.532 - 22.168 | 10.275 | 4.564 - 21.874 | 116 (6.164) | 1.667 | 1.001 - 3.119 | 61 | 31.545 | 16.437 - 99.046 |

[1]Crude data as observed in the study population
[2]Standardized according to gender and municipality
[3]Dark figure - factor: Ratio of standardized seroprevalence (%) and registered positive PCR (%), respectively.
[4]IFR (%): Infection fatality ratio. Percentage of people who have died from or with SARS-CoV-2 infection relative to the calculated number of standardized seropositive individuals
[5]Confidence intervals (CI) were estimated according to Wilson-Score-Method
[6]Confidence intervals (CI) were computed as Bayesian credibility intervals (see methods)

**Supplemental Table 4.** Local seroprevalence (SP) in the study cohort and standardized according to the age and gender distribution of the population of the county Tirschenreuth

| **Local Municipality** | **SP (%)[1]** | **SP (%)[1] 95%-CI[3]** | **SP (%)[2]** | **SP (%)[2] 95%-CI[3]** |
|---|---|---|---|---|
| Bad Neualbenreuth | 7.527 | 3.694 - 14.730 | 7.567 | 3.691 - 14.809 |
| Brand | 5.085 | 1.744 -13.917 | 5.111 | 1.726 - 13.986 |
| Bärnau | 8.466 | 5.278 - 13.308 | 8.510 | 5.291 - 13.382 |
| Ebnath | 4.615 | 1.582 - 12.714 | 4.640 | 1.565 - 12.777 |
| Erbendorf | 7.634 | 4.996 - 11.496 | 7.674 | 5.011 - 11.561 |
| Falkenberg | 2.740 | 0.755 - 9.450 | 2.754 | 0.738 - 9.495 |
| Friedenfels | 5.747 | 2.480 - 12.758 | 5.778 | 2.471- 12.824 |
| Fuchsmühl | 12.644 | 7.209 - 21.238 | 12.711 | 7.221 - 21.354 |
| Immenreuth | 1.000 | 0.177 - 5.449 | 1.005 | 0.164 - 5.471 |
| Kastl | 2.410 | 0.663 - 8.366 | 2.422 | 0.648 - 8.406 |
| Kemnath | 1.701 | 0.729 - 3.919 | 1.710 | 0.726 - 3.940 |
| Konnersreuth | 11.927 | 7.104 - 19.342 | 11.990 | 7.120 - 19.448 |
| Krummennaab | 10.588 | 5.671 - 18.914 | 10.644 | 5.676 - 19.016 |
| Kulmain | 5.147 | 2.515 - 10.243 | 5.174 | 2.513 - 10.299 |
| Leonberg | 18.841 | 11.355 - 29.613 | 18.940 | 11.384 - 29.773 |
| Mitterteich | 18.351 | 14.765 - 22.577 | 18.448 | 14.833 - 22.702 |
| Mähring | 16.822 | 10.914 - 25.031 | 16.911 | 10.949 - 25.168 |
| Neusorg | 6.107 | 3.127 - 11.588 | 6.139 | 3.126 - 11.651 |
| Pechbrunn | 12.500 | 6.718 - 22.081 | 12.566 | 6.724 - 22.200 |
| Plößberg | 6.341 | 3.743 - 10.546 | 6.375 | 3.750 - 10.605 |
| Pullenreuth | 3.636 | 1.423 - 8.979 | 3.656 | 1.414 - 9.025 |
| Reuth b.Erbendorf | 22.535 | 14.375 - 33.515 | 22.654 | 14.420 - 33.696 |
| Tirschenreuth | 9.506 | 7.323 - 12.254 | 9.557 | 7.354 - 12.323 |
| Waldershof | 3.902 | 1.990 - 7.510 | 3.923 | 1.990 - 7.551 |
| Waldsassen | 8.427 | 5.966 - 11.775 | 8.472 | 5.988 - 11.841 |
| Wiesau | 8.261 | 5.352 - 12.541 | 8.305 | 5.368 - 12.611 |

[1]Crude data as observed in the study population
[2]Standardised according to gender and age
[3]Confidence intervals (CI) were estimated according to Wilson-Score-Method.

**Supplemental table 5:** Seroprevalence (SP), dark figure (DF) factor and infection fatality ratio (IFR) in municipality subgroups with and without senior citizen residences in the county of Tirschenreuth

| **Municipality subgroup** | **# in county**<br>**# in study**<br>**# seropositives** | **SP (%)**[1]<br>**95%-CI**[4] | **Registered positive PCR**<br>**#, %** | **DF**[2]<br>**95%-CI**[5] | **Number of deaths** | **IFR (%)**[3]<br>**95%-CI**[5] |
|---|---|---|---|---|---|---|
| with senior citizen residences | 43870<br>2802<br>248 | 8.898<br>7.893 - 10.014 | 888<br>2.024 | 4.083<br>3.828 - 4.989 | 125 | 3.202<br>2.609 - 3.967 |
| without senior citizen residences | 20773<br>1399<br>115 | 8.264<br>6.925- 9.831 | 219<br>1.049 | 8.482<br>6.279 - 9.656 | 13 | 0.754<br>0.436 - 1.334 |

[1]Standardised according to gender and age

[2]Dark figure factor: Ratio of standardized seroprevalence (%) and registered positive PCR (%), respectively.

[3]IFR (%): Infection fatality ratio. Percentage of people who have died from or with CoV-2 infection relative to the calculated number of standardized seropositive individuals

[4]Confidence intervals (CI) were estimated according to Wilson-Score-Method

[5]Confidence intervals (CI) were computed as Bayesian credibility intervals (see methods)

**Supplemental Table 6: Symptoms by serostatus.** Shown are relative (%) and absolute (#) frequencies of reported symptoms by participants per serostatus subgroup (column) with a corresponding [number of participants with non-missing information]. In addition, we show the Odds Ratio, OR, with associated 95%-CI comparing serostatus positive versus serostatus negative (unadjusted; results adjusted for age and sex very similar, data not shown). Participants have been asked whether they had experienced any of the stated symptoms since the start of the pandemic (as per Feb 1st, 2020).

| | **Serostatus positive AND PCR-test pos** | **Serostatus positive AND no PCR-test or neg.** | **Serostatus positive** | **Serostatus negative** | **OR (95%-CI)** |
|---|---|---|---|---|---|
| Coughing, % (#) | 66.7 (46) [n=69] | 41.2 (121) [n=294] | 46.0 (167) [n=363] | 30.0 (1139) [n=3799] | 1.99 [1.6, 2.47] |
| Shortness of Breath. % (#) | 33.3 (23) [n=69] | 10.2 (30) [n=294] | 14.6 (53) [n=363] | 5.8 (220) [n=3799] | 2.78 [2.02, 3.84] |
| Breathing problems, % (#) | 39.1 (27) [n=69] | 18.0 (53) [n=294] | 22.0 (80) [n=363] | 9.2 (351) [n=3799] | 2.78 [2.12, 3.64] |
| Fever, % (#) | 55.1 (38) [n=69] | 30.3 (89) [n=294] | 35.0 (127) [n=363] | 7.6 (289) [n=3799] | 6.54 [5.11, 8.37] |
| Chills, % (#) | 34.8 (24) [n=69] | 20.7 (61) [n=294] | 23.4 (85) [n=363] | 6.5 (248) [n=3799] | 4.38 [3.33, 5.76] |
| Pain in extremities, % (#) | 50.7 (35) [n=69] | 37.1 (109) [n=294] | 39.7 (144) [n=363] | 18.1 (689) [n=3799] | 2.97 [2.37, 3.72] |
| Diarrhea, % (#) | 37.7 (26) [n=69] | 21.8 (64) [n=294] | 24.8 (90) [n=363] | 17.6 (669) [n=3799] | 1.54 [1.2, 1.99] |
| Olfactory problems, % (#) | 58.0 (40) [n=69] | 32.7 (96) [n=294] | 37.5 (136) [n=363] | 2.7 (101) [n=3799] | 21.94 [16.41, 29.32] |
| Taste problems, % (#) | 63.8 (44) [n=69] | 35.4 (104) [n=294] | 40.8 (148) [n=363] | 4.1 (155) [n=3799] | 16.18 [12.43, 21.07] |
| Nausea, % (#) | 17.4 (12) [n=69] | 12.6 (37) [n=294] | 13.5 (49) [n=363] | 8.3 (314) [n=3799] | 1.73 [1.25, 2.39] |
| Eye inflammation, % (#) | 8.7 (6) [n=69] | 4.1 (12) [n=294] | 5.0 (18) [n=363] | 5.3 (200) [n=3799] | 0.94 [0.57, 1.54] |
| Headache, % (#) | 42.0 (29) [n=69] | 35.7 (105) [n=294] | 36.9 (134) [n=363] | 33.1 (1259) [n=3799] | 1.18 [0.94, 1.48] |
| Fatigue, % (#) | 75.4 (52) [n=69] | 53.1 (156) [n=294] | 57.3 (208) [n=363] | 29.1 (1104) [n=3799] | 3.28 [2.63, 4.08] |
| Rhinitis, % (#) | 27.5 (19) [n=69] | 35.0 (103) [n=294] | 33.6 (122) [n=363] | 27.8 (1056) [n=3799] | 1.31 [1.05, 1.65] |
| **Nothing of the above, % (#)** | **2.9 (2) [n=69]** | **15.3 (45) [n=294]** | **12.9 (47) [n=363]** | **35.6 (1353) [n=3799]** | **0.27 [0.2, 0.37]** |

**Supplemental Table 7: Reported symptoms by sex and age groups.** Shown are relative (%) and absolute (#) frequencies of individuals reporting symptoms from a subgroup-specific [number of individuals with non-missing information]. Participants have been asked whether they had experienced any of the stated symptoms since the start of the pandemic (as per Feb 1st, 2020).

| | All | Male | Female | Age 14-19 | Age 20-69 | Age >=70 |
|---|---|---|---|---|---|---|
| Coughing, % (#) | 31.4 (1307) [n=4164] | 32.2 (650) [n=2016] | 30.6 (657) [n=2148] | 37.9 (86) [n=227] | 32.6 (1082) [n=3316] | 22.4 (139) [n=621] |
| Shortness of Breath, % (#) | 6.6 (273) [n=4164] | 5.6 (113) [n=2016] | 7.4 (160) [n=2148] | 3.1 (7) [n=227] | 6.2 (205) [n=3316] | 9.8 (61) [n=621] |
| Breathing problems, % (#) | 10.4 (431) [n=4164] | 10.0 (201) [n=2016] | 10.7 (230) [n=2148] | 6.2 (14) [n=227] | 10.7 (356) [n=3316] | 9.8 (61) [n=621] |
| Fever, % (#) | 10.0 (417) [n=4164] | 9.2 (186) [n=2016] | 10.8 (231) [n=2148] | 15.9 (36) [n=227] | 10.4 (344) [n=3316] | 6.0 (37) [n=621] |
| Chills, % (#) | 8.0 (334) [n=4164] | 6.7 (136) [n=2016] | 9.2 (198) [n=2148] | 7.5 (17) [n=227] | 8.9 (295) [n=3316] | 3.5 (22) [n=621] |
| Pain in extremities, % (#) | 20.0 (834) [n=4164] | 19.1 (386) [n=2016] | 20.9 (448) [n=2148] | 13.2 (30) [n=227] | 20.7 (688) [n=3316] | 18.7 (116) [n=621] |
| Diarrhea, % (#) | 18.2 (759) [n=4164] | 18.1 (364) [n=2016] | 18.4 (395) [n=2148] | 17.2 (39) [n=227] | 19.6 (651) [n=3316] | 11.1 (69) [n=621] |
| Olfactory problems, % (#) | 5.7 (237) [n=4164] | 4.7 (95) [n=2016] | 6.6 (142) [n=2148] | 3.1 (7) [n=227] | 6.1 (202) [n=3316] | 4.5 (28) [n=621] |
| Taste problems, % (#) | 7.3 (303) [n=4164] | 6.5 (132) [n=2016] | 8.0 (171) [n=2148] | 4.0 (9) [n=227] | 8.0 (264) [n=3316] | 4.8 (30) [n=621] |
| Nausea, % (#) | 8.7 (364) [n=4164] | 5.8 (116) [n=2016] | 11.5 (248) [n=2148] | 11.9 (27) [n=227] | 9.3 (307) [n=3316] | 4.8 (30) [n=621] |
| Eye inflammation, % (#) | 5.2 (218) [n=4164] | 4.7 (94) [n=2016] | 5.8 (124) [n=2148] | 2.6 (6) [n=227] | 5.3 (175) [n=3316] | 6.0 (37) [n=621] |
| Headache, % (#) | 33.5 (1394) [n=4164] | 26.6 (536) [n=2016] | 39.9 (858) [n=2148] | 42.7 (97) [n=227] | 37.2 (1232) [n=3316] | 10.5 (65) [n=621] |
| Fatigue, % (#) | 31.5 (1313) [n=4164] | 27.4 (552) [n=2016] | 35.4 (761) [n=2148] | 23.3 (53) [n=227] | 33.7 (1117) [n=3316] | 23.0 (143) [n=621] |
| Rhinitis, % (#) | 28.3 (1179) [n=4164] | 29.9 (602) [n=2016] | 26.9 (577) [n=2148] | 40.5 (92) [n=227] | 30.1 (999) [n=3316] | 14.2 (88) [n=621] |
| Nothing, % (#) | 33.6 (1401) [n=4164] | 35.4 (713) [n=2016] | 32.0 (688) [n=2148] | 27.8 (63) [n=227] | 31.0 (1027) [n=3316] | 50.1 (311) [n=621] |

**Supplemental Table 8: Association of demographic and lifestyle factors with seropositivity.** Shown are Odds Ratios (OR) and 95%-confidence intervals as well as P-values from three logistic regression models. Participants were asked in which profession they were mostly working in February 2020, whether and how much they were smoking and drinking alcohol at the time of the questionnaire completion (from June 19th, 10 days prior to 1st day of the blood draws, until the last day of the blood draws), which was 1-3 weeks before the blood draw to derive serum antibody status. Body-mass-index was derived from self-reported weight and height in the questionnaire and physical activity was assessed as any of category of >= 1 hours per week (medium/high) versus < 1 hours per week (including walking and biking). CI = confidence interval

| | | Model I | | | | Model II | | | | Model III | | | |
|---|---|---|---|---|---|---|---|---|---|---|---|---|---|
| Covariate [reference] | Cat | OR | 95%-CI | | P | OR | 95%-CI | | P | OR | 95%-CI | | P |
| Intercept* | | 0.09 | 0.07 | 0.10 | - | 0.08 | 0.05 | 0.11 | - | 0.09 | 0.05 | 0.14 | - |
| Age [20-69] | 14-19 | 1.20 | 0.75 | 1.84 | 0.422 | 1.20 | 0.72 | 1.92 | 0.463 | 1.24 | 0.72 | 2.05 | 0.427 |
| | 70+ | 0.95 | 0.69 | 1.28 | 0.740 | 1.06 | 0.73 | 1.50 | 0.750 | 0.95 | 0.65 | 1.38 | 0.799 |
| Sex [male] | female | 1.02 | 0.82 | 1.26 | 0.871 | 0.93 | 0.73 | 1.17 | 0.519 | 1.02 | 0.78 | 1.32 | 0.906 |
| Education years [6-10] | 11-15 | - | - | - | - | 1.07 | 0.84 | 1.37 | 0.582 | 0.96 | 0.75 | 1.25 | 0.775 |
| | 16-23 | - | - | - | - | 0.94 | 0.6 | 1.43 | 0.771 | 0.85 | 0.54 | 1.31 | 0.481 |
| Person household [1] | 2 | - | - | - | - | 0.89 | 0.62 | 1.32 | 0.565 | 0.87 | 0.59 | 1.31 | 0.482 |
| | 3-5 | - | - | - | - | 1.05 | 0.73 | 1.55 | 0.793 | 0.99 | 0.67 | 1.49 | 0.963 |
| | 6+ | - | - | - | - | 1.05 | 0.58 | 1.85 | 0.860 | 0.96 | 0.52 | 1.73 | 0.902 |
| Profession [other] | grocery | - | - | - | - | 0.87 | 0.36 | 1.77 | 0.725 | 0.82 | 0.31 | 1.76 | 0.637 |
| | medicine | - | - | - | - | 2.13 | 1.46 | 3.07 | <0.001 | 2.26 | 1.53 | 3.28 | <0.001 |
| Phys. Activity [high] | low | - | - | - | - | - | - | - | - | 0.93 | 0.71 | 1.21 | 0.577 |
| Alcohol drinks / day [0-0.25] | 0 | - | - | - | - | - | - | - | - | 0.76 | 0.52 | 1.09 | 0.144 |
| | 0.25-1 | - | - | - | - | - | - | - | - | 1.28 | 0.95 | 1.72 | 0.101 |
| | >1 | - | - | - | - | - | - | - | - | 1.33 | 0.96 | 1.85 | 0.860 |
| BMI [18.5-25] | <18.5 | - | - | - | - | - | - | - | - | 1.16 | 0.39 | 2.80 | 0.760 |
| | 25-30 | - | - | - | - | - | - | - | - | 1.14 | 0.86 | 1.50 | 0.367 |
| | >30 | - | - | - | - | - | - | - | - | 1.05 | 0.76 | 1.43 | 0.781 |
| Smoking [never] | ex | - | - | - | - | - | - | - | - | 0.97 | 0.74 | 1.27 | 0.843 |
| | current | - | - | - | - | - | - | - | - | 0.36 | 0.24 | 0.53 | <0.001 |

* Intercept corresponds to estimated probability of seropositivity in reference categories

**Follow-up analysis on the smoking finding:**

**Supplemental Table 9:** Odds-ratios and corresponding 95%-CIs (Wald) by age-groups and sex (current smoking vs. serostatus positive), young=14-39, middle age=40-59, old >=60

| **Group** | | **n** | **# pos** | **# smoke** | **% pos. smoke** | **% pos. non-smoke** | **OR** | **95%-CI** |
|---|---|---|---|---|---|---|---|---|
| Overall | | 4176 | 363 | 852 | 4.2 | 9.8 | 0.404 | [0.284, 0.575] |
| Age-group | young | 1332 | 105 | 297 | 4.0 | 9.0 | 0.426 | [0.230, 0.789] |
| | mid | 1476 | 146 | 370 | 4.3 | 11.8 | 0.339 | [0.199, 0.578] |
| | old | 1368 | 112 | 185 | 4.3 | 8.8 | 0.469 | [0.225, 0.979] |
| Sex | male | 2021 | 174 | 441 | 3.4 | 10.1 | 0.315 | [0.183, 0.540] |
| | female | 2155 | 189 | 411 | 5.1 | 9.6 | 0.505 | [0.317, 0.806] |
| Age x Sex | young, male | 650 | 41 | 163 | 3.1 | 7.4 | 0.396 | [0.153, 1.028] |
| | young, female | 682 | 64 | 134 | 5.2 | 10.4 | 0.475 | [0.211, 1.066] |
| | mid, male | 721 | 76 | 185 | 2.7 | 13.2 | 0.182 | [0.072, 0.458] |
| | mid, female | 755 | 70 | 185 | 5.9 | 10.4 | 0.548 | [0.281, 1.066] |
| | old, male | 650 | 57 | 93 | 5.4 | 9.3 | 0.552 | [0.214, 1.420] |
| | old, female | 718 | 55 | 92 | 3.3 | 8.3 | 0.372 | [0.114, 1.217] |

**Supplemental Table 10.** Odds-ratios and corresponding 95%-CIs (Wald) by age-sex group (current smoking vs. PCR-test positive if tested), young=14-39, middle age=40-59, old >=60

| Group | | n | # pos | # smoke | % pos. smoke | % pos. non-smoke | OR | 95%-CI |
|---|---|---|---|---|---|---|---|---|
| Overall | | 501 | 74 | 105 | 6.7 | 16.9 | 0.351 | [0.156, 0.789] |
| Age-group | young | 145 | 16 | 34 | 2.9 | 13.5 | 0.194 | [0.025, 1.525] |
| | mid | 216 | 30 | 58 | 5.2 | 17.1 | 0.265 | [0.077, 0.909] |
| | old | 140 | 28 | 13 | 23.1 | 19.7 | 1.224 | [0.313, 4.780] |
| Sex | male | 190 | 39 | 31 | 9.7 | 22.6 | 0.366 | [0.105, 1.274] |
| | female | 311 | 35 | 74 | 5.4 | 13.1 | 0.38 | [0.129, 1.114] |
| Age x Sex | young, male | 49 | 7 | 11 | 0 | 18.4 | 0 | - |
| | young, female | 96 | 9 | 23 | 4.3 | 11.0 | 0.369 | [0.044, 3.120] |
| | mid, male | 74 | 13 | 14 | 7.1 | 20.0 | 0.308 | [0.037, 2.589] |
| | mid, female | 142 | 17 | 44 | 4.5 | 15.3 | 0.263 | [0.058, 1.206] |
| | old, male | 67 | 19 | 6 | 33.3 | 27.9 | 1.294 | [0.217, 7.730] |
| | old, female | 73 | 9 | 7 | 14.3 | 12.1 | 1.208 | [0.128, 11.377] |

**Supplemental Table 11: Dose-response models for seropositivity versus seronegativity.** Shown are results from logistic regression modeling a linear effect of the number of smoked cigarettes on the binary outcome.

| **All participants (Non-smokers with 0 cigarettes per day)** | | | | | |
|---|---|---|---|---|---|
| Covariate [reference] | Cat | OR | 95%-CI | | P |
| Intercept* | | 0.10 | 0.08 | 0.11 | - |
| Age [20-69] | 14-19 | 1.07 | 0.67 | 1.64 | 0.773 |
| | 70+ | 0.85 | 0.62 | 1.15 | 0.317 |
| Sex [male] | female | 0.99 | 0.80 | 1.23 | 0.926 |
| #Cigs | (per 10) | 0.50 | 0.37 | 0.65 | <0.001 |

| **Current smoker** | | | | | |
|---|---|---|---|---|---|
| Covariate [reference] | Cat | OR | 95%-CI | | P |
| Intercept* | | 0.05 | 0.02 | 0.10 | - |
| Age [20-69] | 14-19 | 1.70 | 0.26 | 6.51 | 0.501 |
| | 70+ | 1.68 | 0.39 | 5.01 | 0.411 |
| Sex [male] | female | 1.46 | 0.74 | 2.93 | 0.281 |
| #Cigs | (per 10) | 0.69 | 0.43 | 1.07 | 0.108 |

* Intercept corresponds to estimated probability of seropositivity in reference categories

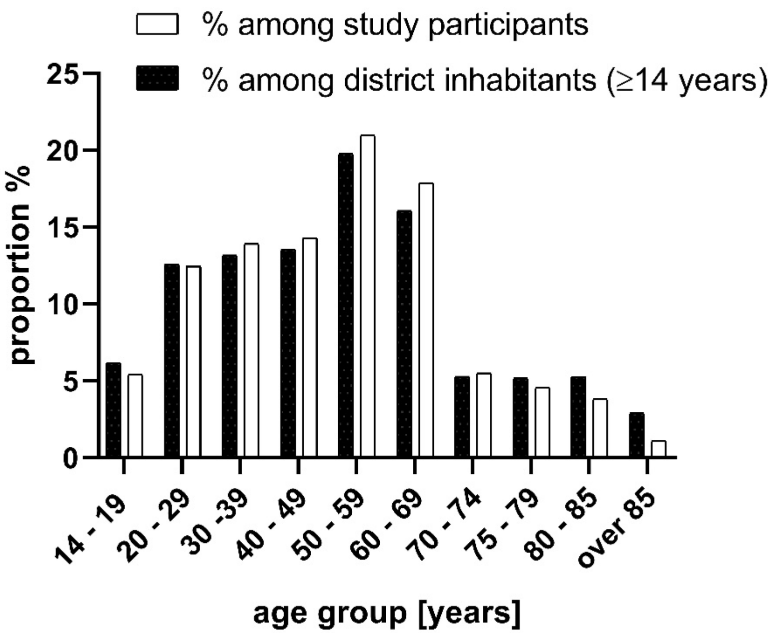


**Supplemental Figure 1**

### Supplemental Figure 2

**Nonlinear effect number of cigarettes**

**All (p<0.001)**

**Smoker (p=0.086)**

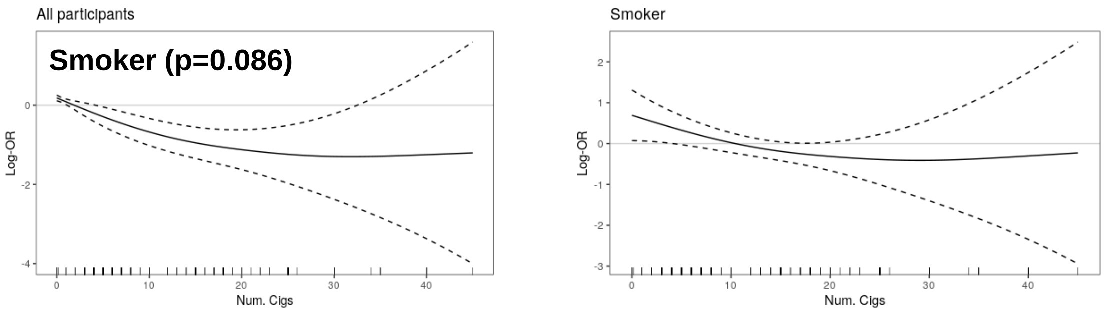

**Legends to Supplemental Figures**

**Supplemental Figure 1** Proportion (%) of TiKoCo-19 study participants versus county population in the various age groups.

*Figure was designed using GraphPad Prism version 8.4.3 for Windows, GraphPad Software, La Jolla California USA, www.graphpad.com*

**Supplemental Figure 2** Non-linear association of number of cigarettes with seropositivity. Log-OR are shown for all participants and active smokers. P-values are indicated.

*Analysis was conducted in R (R Core Team (2020). R: A language and environment for statistical computing. R Foundation for Statistical Computing, Vienna, Austria. URL http://www.R-project.org/) and figure 2 was produced using the package ggplot2 (Wickham, H. (2009)* ggplot2: elegant graphics for data analysis*. Springer New York).*
